# Sex and genotype influence the disruption of circulating NMDAR-related amino acids in patients with Parkinson’s disease

**DOI:** 10.1101/2025.08.08.25333287

**Authors:** Isar Yahyavi, Federica Carrillo, Tommaso Nuzzo, Matteo Vidali, Sara Pietracupa, Nicola Modugno, Anna Di Maio, Francesco Errico, Teresa Esposito, Alessandro Usiello

**Author notes:** to whom correspondence should be addressed Corresponding authors: Dr Teresa Esposito PhD Molecular Genetics and Genomics Laboratory Institute of Genetics and Biophysics, “Adriano Buzzati Traverso”, Italian National Research Council (CNR) 80131 Naples – Italy. PI IGB-CNR laboratory IRCCS INM Neuromed 86077 Pozzilli – Italy., Prof Alessandro Usiello PhD. Department of Environmental, Biological and Pharmaceutical Sciences and Technologies (DISTABIF). Università della Campania, L. Vanvitelli, Viale Abramo Lincoln 5, 81100, Caserta, Italy. PI Neuroscience Laboratory. CEINGE, Biotecnologie Avanzate Franco Salvatore, S.c.a.rl. 80145 Naples-Italy. contributed equally and share joint first authorship (Isar Yahyavi, Federica Carrillo, Tommaso Nuzzo).

## Abstract

Sex and genetic factors influence the epidemiology and clinical features of Parkinson’s disease (PD), but their impact on N-methyl-D-aspartate receptor (NMDAR)-related amino acids remains unclear. We measured serum levels of these molecules by high-performance liquid chromatography (HPLC) in a well-characterized cohort of PD patients (n = 245) and healthy controls (HCs, n = 203), stratified by sex and disease subtype (idiopathic, n = 121; genetic, n = 124). The genetic subgroup included carriers of pathogenic variants in *LRRK2*, *TMEM175*, *PARK2*, *PINK1*, *PARK7*, and *GBA1*. We identified marked sex-dependent differences in amino acid profiles, confirmed by linear models with interaction terms. Idiopathic male patients showed reductions in serum levels of L-glutamate, L-glutamine, L-aspartate, D-serine, L-serine, glycine, and L-asparagine compared with HCs, whereas genetic PD males displayed more limited changes. Female patients showed minimal or no significant alterations regardless of subtype. Within the PD cohort, idiopathic males exhibited lower levels of L-glutamate, L-aspartate, glycine, D-serine, and L-serine than genetic PD males, while no differences emerged between female subgroups. These findings reveal a previously unrecognized sex-and genotype-dependent regulation of NMDAR-related amino acid homeostasis in PD, highlighting the importance of biological stratification for biomarker discovery and for developing targeted, personalized therapeutic strategies.

## Introduction

Parkinson’s disease (PD) is the second most common neurodegenerative disorder, characterized by the progressive loss of dopaminergic neurons in the substantia nigra pars compacta combined with the accumulation of α-synuclein aggregates forming Lewy bodies ^1,2^. Clinically, PD manifests with both motor symptoms, including tremors at rest, bradykinesia, rigidity, and postural instability, and non-motor symptoms, such as constipation, depression, anosmia, and sleep disorders, which often precede motor onset. In later stages, cognitive impairment and dysautonomia may also emerge^2,3^.

Beyond dopaminergic neurodegeneration, increasing evidence indicates that glutamatergic dysfunction plays a key role in PD pathophysiology ^4,5^. In particular, alterations in N-methyl-D-aspartate receptor (NMDAR) signaling within the basal ganglia have been implicated in synaptic plasticity defects, L-DOPA-induced dyskinesia and cognitive deficits^6–12^.

NMDAR activity is primarily driven by L-glutamate (L-Glu) and L-aspartate (L-Asp), the main excitatory neurotransmitters in the central nervous system (CNS)^13,14^. In addition, D-serine (D-Ser) and glycine (Gly) act as endogenous co-agonists of NMDARs, while L-serine (L-Ser) serves as their precursor^15–17^. Disruption of NMDAR-related amino acid homeostasis has been reported across central and peripheral tissues, including post-mortem caudate putamen, cerebrospinal fluid and serum in PD patients and MPTP-treated monkeys^18–21^. Consistent with this evidence, experimental and clinical studies have shown that stimulation of the NMDAR glycine-binding site enhances striatal dopaminergic reinnervation and improves motor and non-motor symptoms in PD patients and animal models^22–28^.

In recent years, PD has been increasingly recognized as a multisystem disorder rather than a brain-centered motor disease^29,30^. Within this framework, PD is now understood as a complex and heterogenous condition in which sex and genetic background influence disease risk, progression, clinical manifestations, and treatment response^31,32^. In this scenario, multiple studies have reported alterations in amino acid and lipid metabolism in PD^33–43^. However, it remains unclear whether dysregulation of circulating NMDAR-related amino acids varies according to sex and PD subtype.

To address this gap, we analyzed serum levels of NMDAR-related amino acids and their precursors—specifically L-Glu, L-glutamine (L-Gln), L-Asp, L-asparagine (L-Asn), L-Ser, D-Ser, and Gly—using high-performance liquid chromatography (HPLC) in a well-characterized cohort of PD patients stratified by sex and disease subtype (idiopathic and genetic), alongside sex-matched healthy controls. We further assessed the relationship between amino acid levels and demographic and clinical variables, including age, disease duration, age at onset, Levodopa Equivalent Daily Dose (LEDD), and motor symptom severity (MDS-UPDRS III).

## Results

### Clinical and demographic characteristics of the study cohort

We analyzed demographic and clinical data from 245 PD patients and 203 HCs **(Table 1)**. PD patients were significantly older and included a higher proportion of males compared to HCs **(Table 1)**. Within the PD cohort, 121 patients were classified as idiopathic PD (iPD; 65 males, 56 females) and 124 as genetic PD (gPD; 64 males, 60 females), based on the presence of pathogenic mutations **(Table 2**; **Table S1**).

**Table 1.**
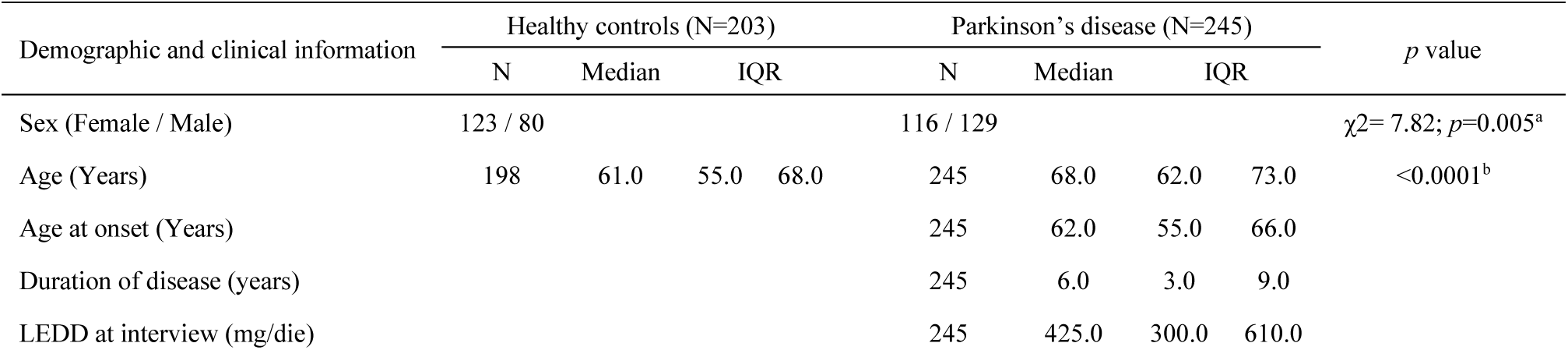
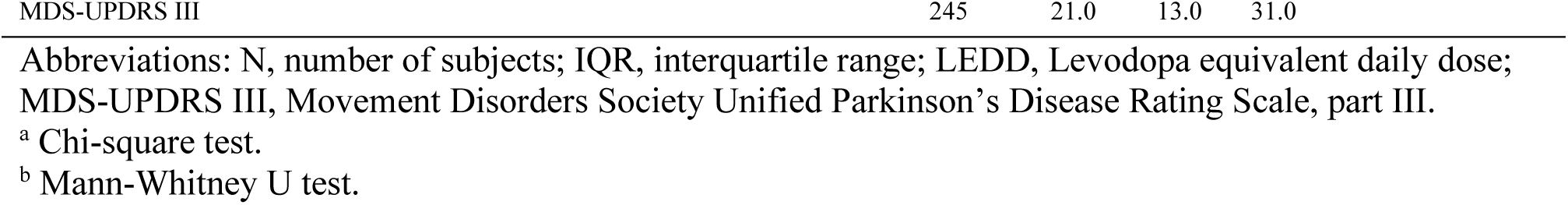
Demographic and clinical features of Parkinson’s disease patients and healthy controls subjects.

**Table 2.** Demographic and clinical features of idiopathic PD, genetic PD and HC groups.

| Demographic and clinical information | Healthy controls (N=203) |  |  | Idiopathic PD (N=121) |  |  | Genetic PD (N=124) |  |  | *p* value |
| --- | --- | --- | --- | --- | --- | --- | --- | --- | --- | --- |
|  | N | Median | IQR | N | Median | IQR | N | Median | IQR |  |
| Sex (Female / Male) | 123 / 80 | | | 56 / 65 | | | 60 / 64 | | | $p = 0.019$; $\chi^2 = 7.933^a$ |
| Age (years) | 198 | 61.0 | 55.0 68.0 | 121 | 68*** | 62.0 74.0 | 124 | 68*** | 62.5 72.5 | $<0.0001^b$ |
| Age at onset (years) |  |  |  | 121 | 62.0 | 55.0 68.0 | 124 | 61.0 | 55.0 65.0 | 0.123<sup>c</sup> |
| Duration of disease (years) |  |  |  | 121 | 5.0 | 3.0 8.0 | 124 | 6.0 | 3.0 10.0 | 0.03<sup>c</sup> |
| LEDD at interview (mg/die) |  |  |  | 121 | 400.0 | 300.0 580.0 | 124 | 457.5 | 305.0 675.0 | 0.322<sup>c</sup> |
| MDS-UPDRS III |  |  |  | 121 | 21.0 | 13.0 32.0 | 124 | 19.0 | 13.0 31.0 | 0.453<sup>c</sup> |

Abbreviations: N, number of subjects; IQR, interquartile range; HC, healthy controls; LEDD, Levodopa equivalent daily dose; MDS-UPDRS III, Movement Disorders Society Unified Parkinson's Disease Rating Scale, part III.
<sup>a</sup> Chi-square test.
<sup>b</sup> Kruskal-Wallis test.
<sup>c</sup> Mann-Whitney U test.
\*\*\* $p < 0.0001$ , compared to HC (Dunn's multiple comparisons test)

The two subgroups differed from HCs in terms of age and sex distribution, while they were comparable in age, age at onset, LEDD, and motor symptom scores (MDS-UPDRS III) **(Table 2)**. Notably, gPD patients had longer disease duration, compared to the iPD group **(Table 2)**. Sex-stratified analyses confirmed comparable clinical features between iPD and gPD males (**Table S2**). In contrast, gPD females showed earlier disease onset and longer motor symptoms duration than iPD females (**Table S2**). No significant differences were observed among carriers of different pathogenic mutations.

### PD patients exhibit alterations in circulating NMDAR-related amino acids at the whole-cohort level

To evaluate the influence of PD on circulating NMDAR-related amino acids, we measured serum levels of D-Ser, L-Ser, Gly, L-Glu, L-Gln, L-Asp, and L-Asn in PD patients and HCs by HPLC (**Figure 1a; Table S3**). We also calculated the D-Ser/total Ser and L-Gln/L-Glu ratios as indices of the conversion of D-Ser and L-Glu from their respective precursors, L-Ser and L-Gln.

**Figure 1.**
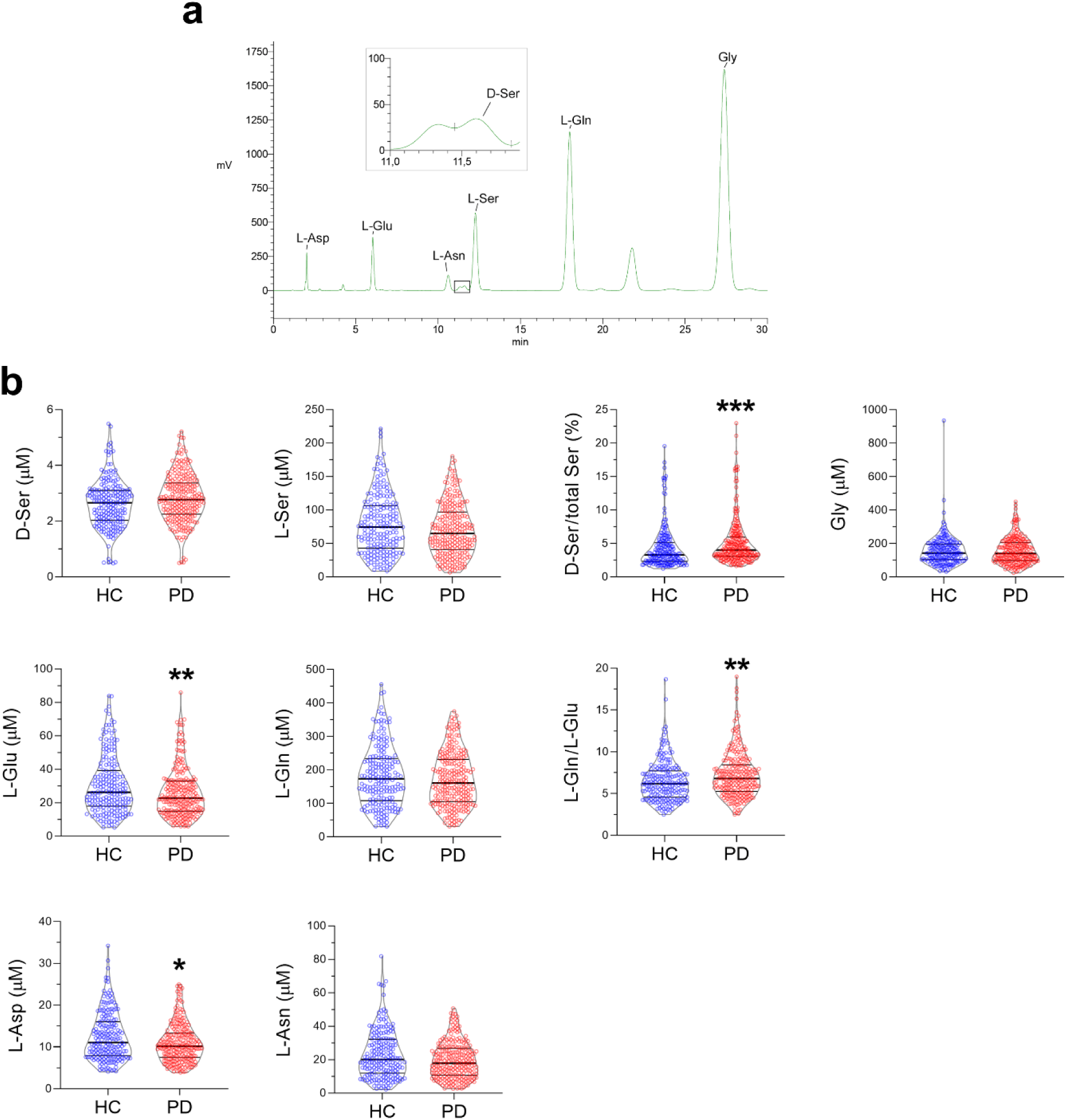
D-and L-amino acids levels in the serum of Parkinson‘s disease patients and healthy subjects. **(a)** Representative HPLC chromatogram illustrating the separation and detection of D-serine (D-Ser), L-serine (L-Ser), glycine (Gly), L-glutamate (L-Glu), L-glutamine (L-Gln), L-aspartate (L-Asp), and L-asparagine (L-Asn) peaks from a blood serum sample. The magnification of D-Ser peak is shown. **(b)** Violin plots representing D-Ser, L-Ser, D-Ser/total Ser ratio, Gly, L-Glu, L-Gln, L-Gln/L-Glu ratio, L-Asp, and L-Asn levels in the serum of Parkinson’s disease patients (PD, N=245) compared to healthy control subjects (HC, N=203). The amino acid content is expressed as µM. In each sample, free amino acids were detected in a single run. Dots represent the single subjects’ values, while lines illustrate the median with interquartile range. \**p* < 0.05, \*\**p* < 0.01, \*\*\**p* < 0.001 (Mann–Whitney U test). Only significant differences confirmed by age-, sex-, and LEDD-adjusted ANCOVA are displayed.

Non-parametric Mann-Whitney analysis revealed significantly elevated serum D-Ser levels and D-Ser/total Ser ratio, along with reduced L-Glu, L-Asp, and L-Asn levels in PD patients compared with HCs (**Figure 1b; Table S3**). A mild increase in the L-Gln/L-Glu ratio was also observed in the PD group (**Figure 1b; Table S3**). Variations in L-Glu, L-Asp, D-Ser/total Ser and L-Gln/L-Glu ratios remained significant after adjustment for age, sex, and LEDD (ANCOVA analysis on natural log-transformed data) (**Figure 1b; Table S3**).

Notably, these alterations appeared relatively limited at the whole-cohort level, suggesting underlying heterogeneity within the PD population.

### Sex stratification reveals marked reduction of NMDAR-related amino acids in male PD patients

To assess sex-specific differences, we compared male and female PD patients with sex-matched HCs **(Figure 2)**. Remarkably, male patients showed a broad reduction in multiple amino acids, including L-Ser, Gly, L-Glu, L-Gln, L-Asp and L-Asn, together with an increased D-Ser/total Ser ratio **(Figure 2a; Table S4)**. These differences remained significant after adjustment for age and LEDD **(Figure 2a; Table S4)**.

**Figure 2.**
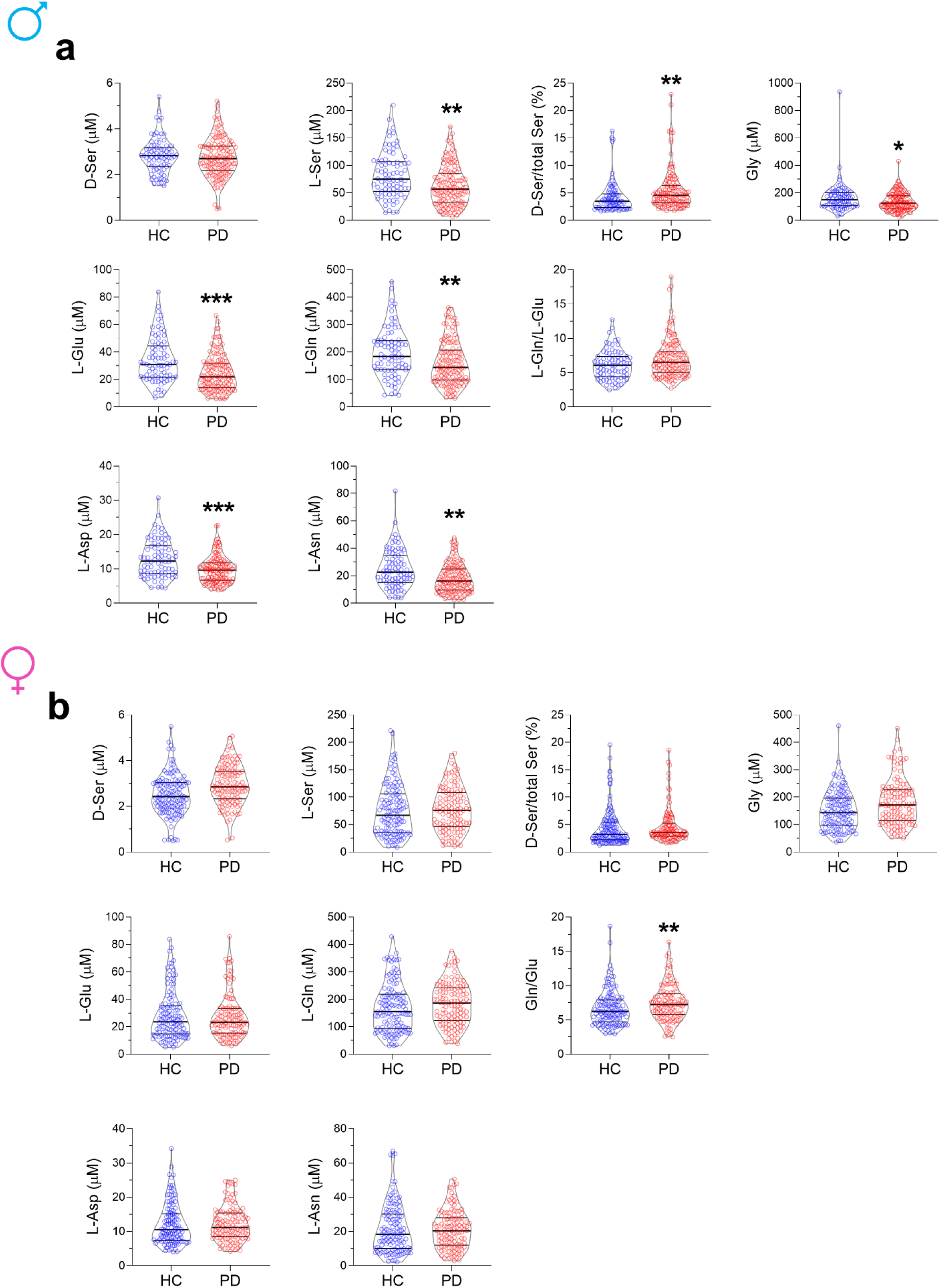
Serum D-and L-amino acids content in male and female Parkinson‘s disease patients and healthy subjects. Analysis of D-serine (D-Ser), L-serine (L-Ser), D-Ser/total Ser ratio, glycine (Gly), L-glutamate (L-Glu), L-glutamine (L-Gln), L-Gln/L-Glu ratio, L-aspartate (L-Asp), and L-asparagine (L-Asn) levels in the serum of male **(a)** and female **(b)** Parkinson’s disease patients (male, N=129; female, N=116) compared to healthy control (HC) subjects (male, N=80; female N=123). The amino acid content is expressed as µM. In each sample, free amino acids were detected in a single run. Dots represent the single subjects’ values, while lines illustrate the median with interquartile range. \**p* < 0.05, \*\**p* < 0.01, \*\*\**p* < 0.001 (Mann–Whitney U test). Only significant differences confirmed by age-and LEDD-adjusted ANCOVA are displayed.

In contrast, female PD patients showed only limited changes, with increased D-Ser, Gly and L-Gln/L-Glu ratio, compared to sex-matched HCs (**Figure 2b; Table S4**). However, only the L-Gln/L-Glu ratio remained significant after adjustment (**Figure 2b; Table S4**).

These findings indicate that amino acid dysregulation in PD is primarily driven by male patients, whereas females show minimal changes.

### Idiopathic PD drives most NMDAR-related amino acid alterations

To evaluate the impact of genetic background, we stratified PD patients into iPD and gPD subgroups. gPD patients carried at least one pathogenic variant in common PD-associated genes, such as *LRRK2*, *TMEM175*, *PARK2*, *PINK1*, *PARK7*, and *GBA1* **(Table 2; Table S1**).

iPD patients showed significant reductions in multiple amino acids, including L-Ser, L-Glu, L-Asp and L-Asn, and an elevated L-Gln/L-Glu ratio, compared to HCs (**Figure 3; Table S5**). These variations remained significant after correction for age, sex, and LEDD (**Table S5)**. In contrast, gPD patients displayed minimal alterations, limited to increased D-Ser levels **(Figure 3; Table S5)**.

**Figure 3.**
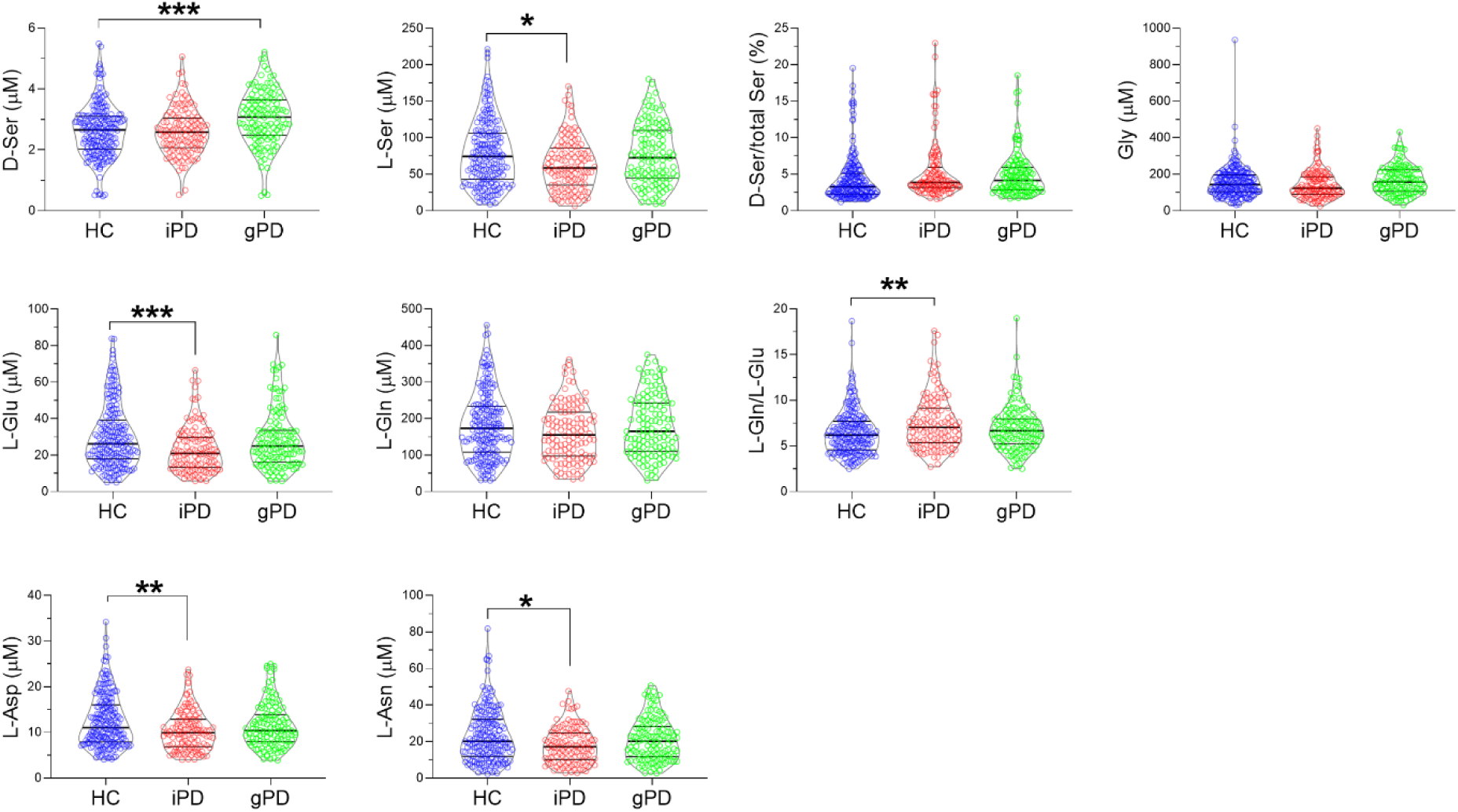
Serum D-and L-amino acids profile in idiopathic PD, genetic PD and healthy controls. Analysis of D-serine (D-Ser), L-serine (L-Ser), D-Ser/total Ser ratio, glycine (Gly), L-glutamate (L-Glu), L-glutamine (L-Gln), L-Gln/L-Glu ratio, L-aspartate (L-Asp), and L-asparagine (L-Asn) levels in the serum of idiopathic PD (iPD, N=121), genetic PD (gPD, N=124) and healthy controls (HC, N=203). The amino acid content is expressed as µM. In each sample, free amino acids were detected in a single run. Dots represent the single subjects’ values, while lines illustrate the median with inter quartile. \**p* < 0.05, \*\**p* < 0.01, \*\*\**p* < 0.001 (post hoc Dunn’s test). Significant post hoc results are shown when Kruskal–Wallis test was confirmed by age-, sex-, and LEDD-adjusted ANCOVA.

Direct comparison between iPD and gPD subgroups confirmed lower levels of several amino acids (L-Ser, D-Ser, Gly and L-Glu) in iPD compared to gPD patients (**Figure S1**). Overall, our findings indicate that amino acid downregulation in PD patients is driven by individuals with idiopathic conditions.

### Sex and PD subtype stratification shows a selective reduction of NMDAR-related amino acids in idiopathic male patients

To further dissect the combined effects of sex and genotype, we analyzed amino acid levels within each sex group across iPD, gPD and HCs.

Combined stratification showed that iPD males exhibited the most pronounced alterations, with broad reductions in D-Ser, L-Ser, Gly, L-Glu, L-Gln, L-Asp, and L-Asn compared to controls (**Figure 4a; Table S6**). In contrast, gPD males showed only a reduction in L-Asp (**Figure 4a; Table S6)**. Both subgroups displayed an increased D-Ser/total Ser ratio compared to HCs (**Figure 4a; Table S6**).

**Figure 4.**
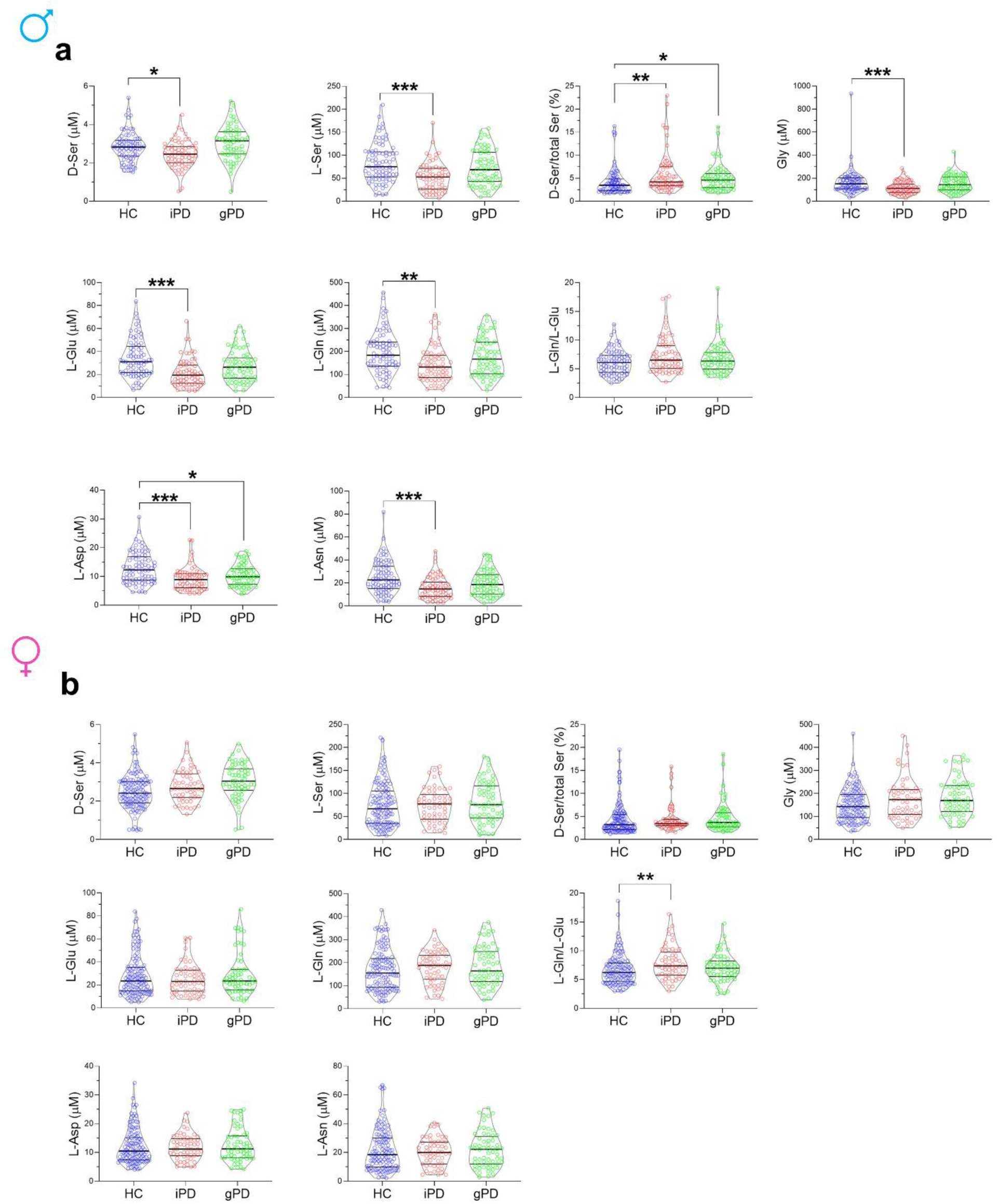
Serum D-and L-amino acids content in male and female subgroups of idiopathic PD, genetic PD and healthy controls. Analysis of D-serine (D-Ser), L-serine (L-Ser), D-Ser/total Ser ratio, glycine (Gly), L-glutamate (L-Glu), L-glutamine (L-Gln), L-Gln/L-Glu ratio, L-aspartate (L-Asp), and L-asparagine (L-Asn) levels in the serum of male **(a)** and female **(b)** idiopathic PD (iPD; male N=65, female N=56), genetic PD (gPD; male N= 64, female N= 60) and healthy controls (HC, male N=80, female N=123). The amino acid content is expressed as µM. In each sample, free amino acids were detected in a single run. Dots represent the single subjects’ values, while lines illustrate the median with interquartile range. \**p* < 0.05, \*\**p* < 0.01, \*\*\**p* < 0.001 (post hoc Dunn’s test). Significant post hoc results are shown when Kruskal–Wallis test was confirmed by age-, and LEDD-adjusted ANCOVA.

In contrast, female patients showed minor alterations compared with HCs, with only the L-Gln/L-Glu ratio remaining significantly altered in iPD females after adjustment for age and LEDD (**Figure 4b; Table S7**).

Direct comparisons confirmed significantly lower levels of D-Ser, L-Ser, Gly, L-Glu, L-Gln, L-Asp, and L-Asn in iPD males compared to gPD males **(Figure 5a; Table S8)**, whereas no consistent differences were observed in females **(Figure 5b; Table S8)**. These findings highlight male iPD patients as the PD subgroup with the most pronounced NMDAR-related amino acid dysregulation.

**Figure 5.**
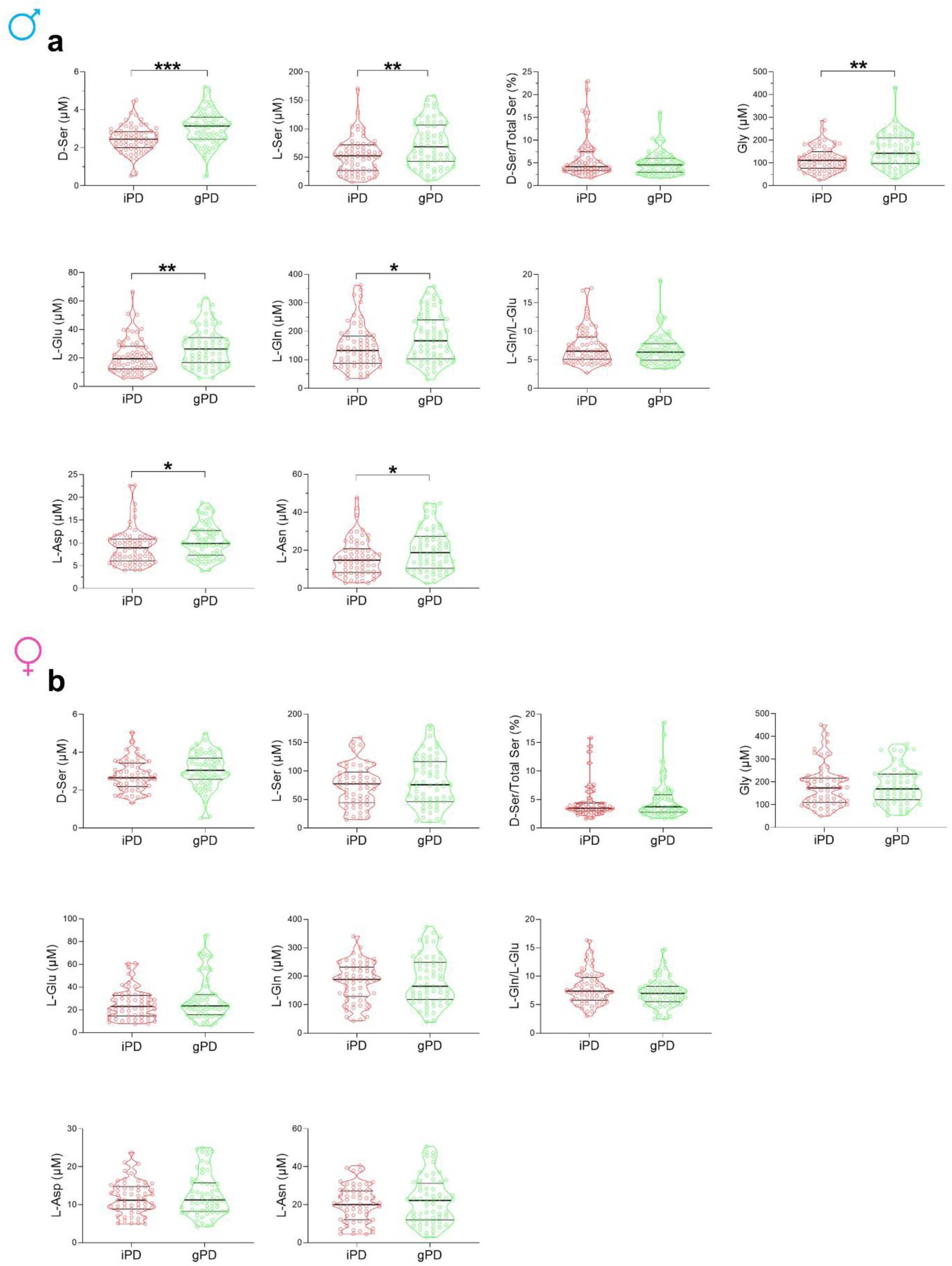
Comparison of serum D-and L-amino acid levels in male and female Parkinson’s disease patients compared between idiopathic and genetic forms. Analysis of D-serine (D-Ser), L-serine (L-Ser), D-ser/total Ser ratio, glycine (Gly), L-glutamate (L-Glu), L-glutamine (L-Gln), L-Gln/L-Glu ratio, L-aspartate (L-Asp), and L-asparagine (L-Asn) in the serum of male **(a)** and female **(b)** idiopathic PD (iPD; male N=65, female N=56) and genetic PD (gPD; male N= 64, female N=60). The amino acid content is expressed as µM. In each sample, free amino acids were detected in a single run. Dots represent the single subjects’ values, while lines illustrate the median with interquartile range. \**p* < 0.05, \*\**p* < 0.01, \*\*\**p* < 0.001 (Mann–Whitney U test). Only differences confirmed by age-, LEDD-and disease duration-adjusted ANCOVA are displayed.

### Integrated modelling confirms sex as the primary driver of amino acid dysregulation

To formally assess whether the effect of PD diagnosis on serum amino acid levels differed by sex, we fitted linear models including a group × sex interaction term, adjusting for age (**Table S9**).

In the full cohort, the interaction was significant for several amino acids, including L-Asp, L-Glu, L-Asn, D-Ser, L-Ser, L-Gln, and Gly, after FDR correction (all p_FDR < 0.01), indicating that PD-associated reductions were significantly more pronounced in males, as shown by the negative direction of the interaction coefficient **(Figure 6)**. These findings confirm that the effect of PD differs between males and females, as reflected in the sex-stratified analyses reported above **(Figure 2, Table S9)**.

**Figure 6.**
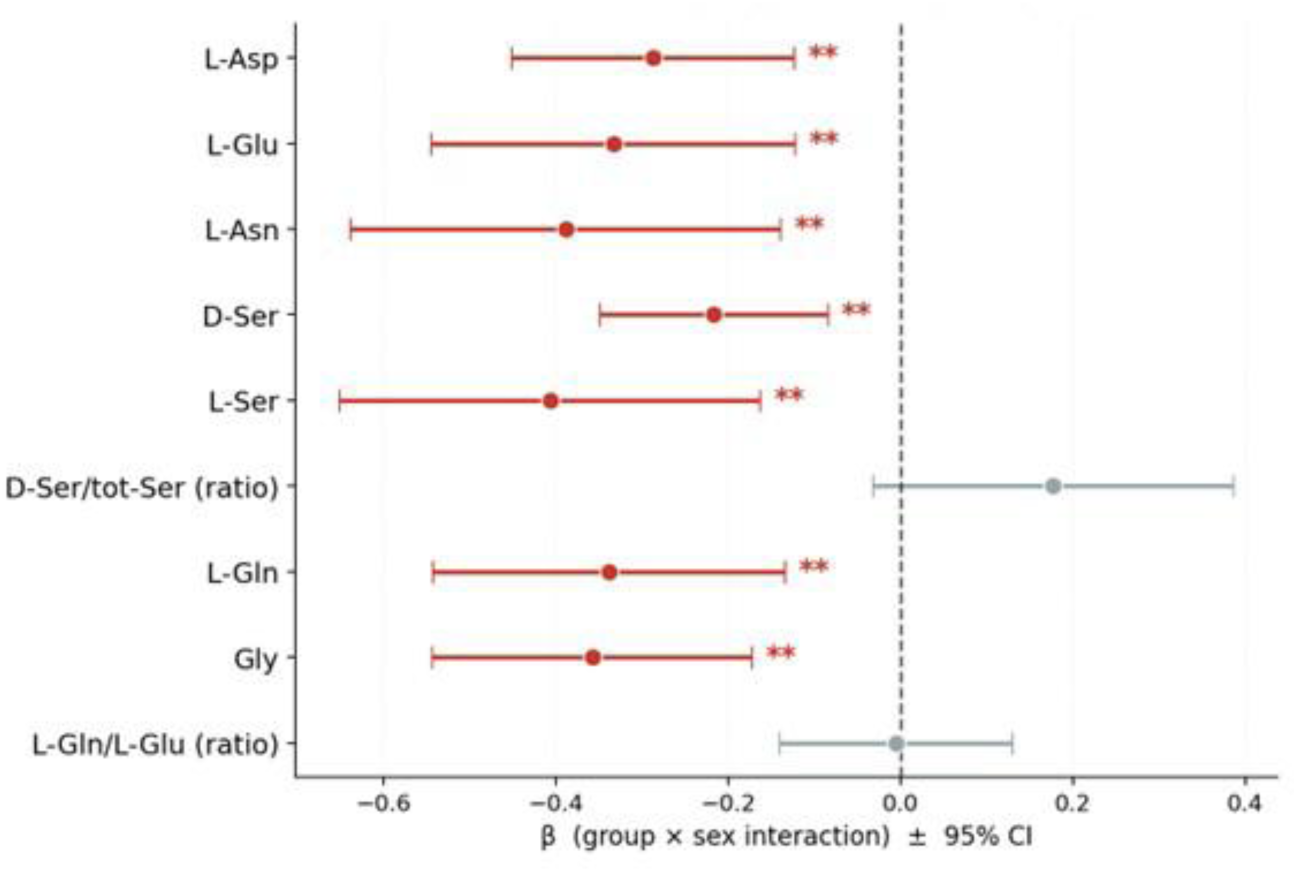
Formal test of group × sex interaction on serum NMDAR-related amino acid levels. Forest plot showing the β coefficients of the group × sex interaction term from linear models fitted separately for each amino acid outcome: log(amino acid) ∼ group + sex + age + group:sex, where group = HC (0) vs PD (1) and sex = female (0) vs male (1). A negative β indicates that the reduction in serum levels associated with PD diagnosis is significantly greater in males than in females. Error bars represent 95% confidence intervals based on heteroscedasticity-consistent (HC3) robust standard errors. Asterisks denote significance after Benjamini-Hochberg false discovery rate (FDR) correction for multiple comparisons (9 tests): **p_FDR < 0.01. Red = significant after FDR correction; grey = not significant.

Given that male PD patients exhibited the most pronounced amino acid dysregulation, we next examined whether this sex-specific pattern was further modulated by disease subtype. To this end, we fitted a linear model restricted to PD patients, including a subtype × sex interaction, adjusting for age, disease duration, and LEDD **(Tables S10 and S11; Figure S2)**.

The subtype × sex interaction did not reach significance for any amino acid after FDR correction (all p_FDR > 0.32), indicating that the relative difference between iPD and gPD does not depend on sex.

In contrast, the main effect of sex was significant for multiple amino acid outcomes, including L-Asp, L-Ser, L-Gln, Gly, L-Asn, and D-Ser, confirming that male PD patients exhibit lower serum levels than females, independently of disease subtype **(Tables S10 and S11)**.

To further examine subtype-specific effects within the male subgroup, we refitted the model using males as the reference category. In this analysis, the main effect of subtype reflects the difference between iPD and gPD in males. This effect was significant after FDR correction for L-Glu, L-Asn, D-Ser, L-Ser, and Gly (all p_FDR < 0.05), with lower levels observed in iPD compared to gPD patients **(Tables S10 and S11)**. No significant subtype effects were observed for females.

Overall, linear models further support that sex is the primary determinant of serum NMDAR-related amino acid variations in PD, while disease subtype contributes to a lesser extent.

### Sex-and subtype-specific correlations between amino acid levels and demographic and clinical variables

We investigated the relationship between amino acid levels and demographic or clinical variables across sexes and PD subtypes **(Table 2; Table S12)**.

Spearman’s correlation analysis revealed sex-specific patterns. Indeed, in females but not in males, D-Ser levels and the D-Ser/total Ser ratio showed positive correlation with age in HCs and iPD patients, whereas no association was observed in gPD patients **(Table 3; Figure S3a,b)**.

**Table 3.** Correlations analysis between serum levels of D-and L-amino acids with age of idiopathic PD, genetic PD and healthy individuals.

| Amino acids | Spearman's correlation | Male |  |  | Female |  |  |
| --- | --- | --- | --- | --- | --- | --- | --- |
|  |  | Healthy controls | Idiopathic PD | Genetic PD | Healthy controls | Idiopathic PD | Genetic PD |
|  |  | Age (years) | Age (years) | Age (years) | Age (years) | Age (years) | Age (years) |
| L-aspartate (μM) | R | 0.003 | -0.068 | 0.112 | 0.008 | 0.190 | -0.071 |
|  | *p* value | 0.981 | 0.588 | 0.377 | 0.929 | 0.160 | 0.590 |
|  | N | 77 | 65 | 64 | 121 | 56 | 60 |
| L-glutamate (μM) | R | 0.022 | -0.044 | 0.128 | 0.143 | 0.198 | -0.096 |
|  | *p* value | 0.851 | 0.731 | 0.314 | 0.118 | 0.144 | 0.465 |
|  | N | 77 | 65 | 64 | 121 | 56 | 60 |
| L-asparagine (μM) | R | 0.001 | -0.038 | 0.144 | -0.032 | 0.208 | -0.076 |
|  | *p* value | 0.995 | 0.762 | 0.257 | 0.731 | 0.124 | 0.562 |
|  | N | 77 | 65 | 64 | 121 | 56 | 60 |
| D-serine (μM) | R | 0.119 | -0.042 | 0.070 | 0.183 | 0.38 | 0.189 |
|  | *p* value | 0.301 | 0.738 | 0.584 | **0.044** | **0.004** | 0.148 |
|  | N | 77 | 65 | 64 | 121 | 56 | 60 |
| L-serine (μM) | R | -0.05 | -0.023 | 0.136 | -0.038 | 0.148 | -0.032 |
|  | *p* value | 0.668 | 0.853 | 0.286 | 0.676 | 0.278 | 0.808 |
|  | N | 77 | 65 | 64 | 121 | 56 | 60 |
| D-serine/total serine (%) | R | 0.102 | 0.039 | -0.121 | 0.218 | 0.053 | 0.181 |
|  | *p* value | 0.376 | 0.760 | 0.342 | **0.016** | 0.699 | 0.167 |
|  | N | 77 | 65 | 64 | 121 | 56 | 60 |
| L-glutamine (μM) | R | -0.02 | -0.010 | 0.125 | -0.002 | 0.237 | -0.044 |
|  | *p* value | 0.861 | 0.934 | 0.326 | 0.983 | 0.079 | 0.737 |
|  | N | 77 | 65 | 64 | 121 | 56 | 60 |
| Glycine (μM) | R | -0.074 | -0.052 | 0.081 | 0.051 | 0.157 | 0.001 |
|  | *p* value | 0.525 | 0.684 | 0.524 | 0.576 | 0.248 | 0.996 |
|  | N | 77 | 65 | 64 | 121 | 56 | 60 |
| L-Glutamine/L-Glutamate | R | -0.011 | -0.002 | -0.060 | -0.195 | -0.139 | 0.158 |
|  | *p* value | 0.924 | 0.989 | 0.640 | **0.032** | 0.307 | 0.227 |
|  | N | 77 | 65 | 64 | 121 | 56 | 60 |

Abbreviations: N, number of subjects. Significant *p* values are shown in bold.

Regarding clinical variables, no significant correlations were found in male patients **(Table 4; Figure S4a)**. In contrast, in females, D-Ser levels showed a positive correlation with age at onset in the iPD group **(Table 4; Figure S4b)**, consistent with a previous finding ^37^.

**Table 4.**
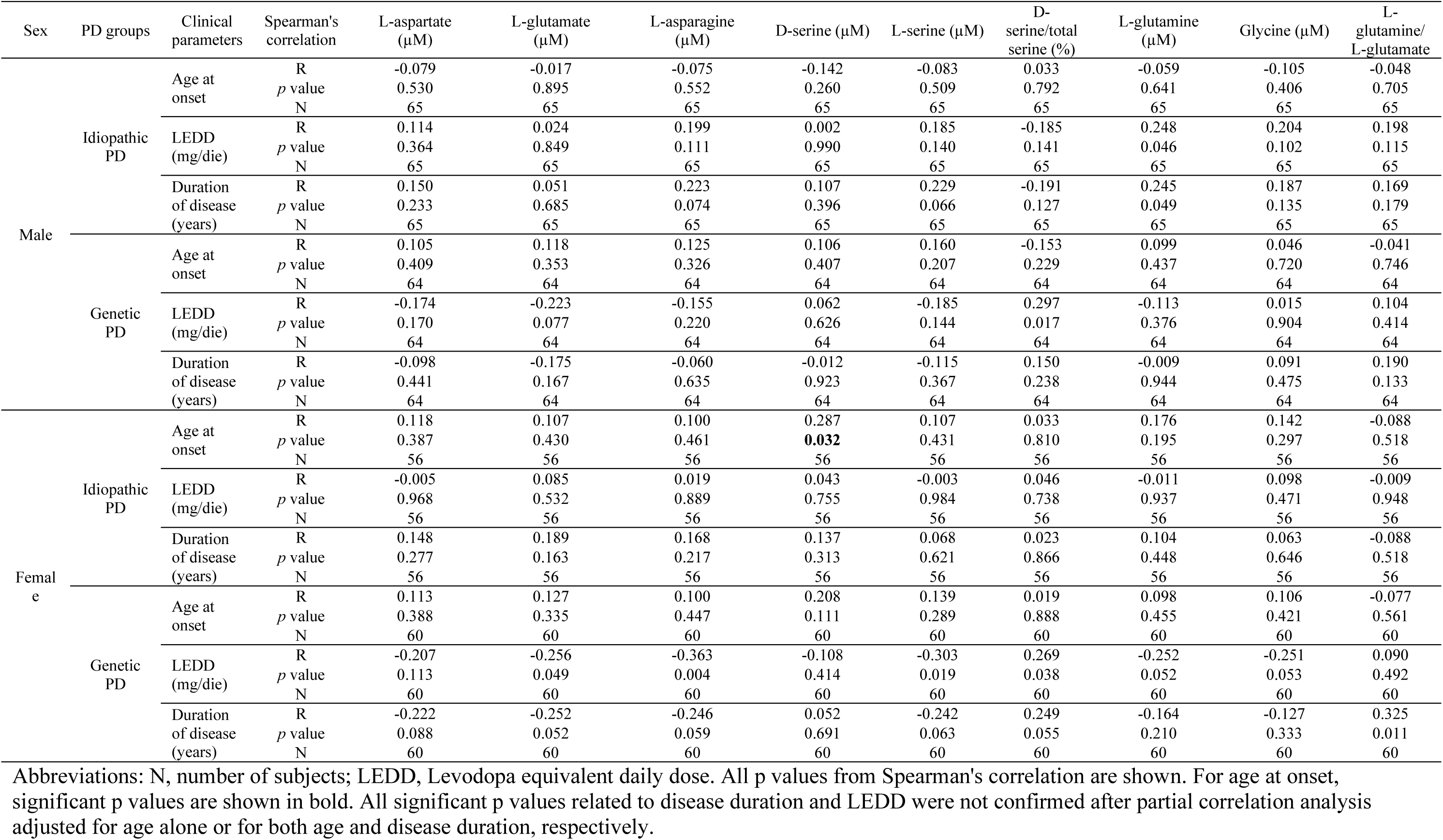
Correlation analysis of serum D- and L-amino acids level with age of disease onset, LEDD and disease duration of PD groups.

Overall, our results indicate that the relationship between D-Ser levels and demographic or clinical features is modulated by both sex and PD subtype.

### Serum D-serine levels correlate with motor symptom severity in patients with genetic Parkinson’s disease

We next investigated the correlation between amino acid levels and motor symptom severity (MDS-UPDRS III scores) **(Table 5)**.

**Table 5.** Correlation analysis of serum D- and L- amino acids levels with MDS-UPDRS III of patients with idiopathic and genetic PD.

|  | Sex | PD groups | Spearman's correlation | L-aspartate (μM) | L-glutamate (μM) | L-asparagine (μM) | D-serine (μM) | L-serine (μM) | D-serine/total serine (%) | L-glutamine (μM) | Glycine (μM) | L-glutamine / L-glutamate |
| --- | --- | --- | --- | --- | --- | --- | --- | --- | --- | --- | --- | --- |
| MDS-UPDRS III | Male | Idiopathic PD | R | 0.036 | -0.060 | 0.102 | 0.026 | 0.110 | -0.106 | 0.109 | 0.149 | 0.146 |
|  |  |  | *p* value | 0.774 | 0.636 | 0.419 | 0.839 | 0.384 | 0.402 | 0.386 | 0.237 | 0.247 |
|  |  |  | N | 65 | 65 | 65 | 65 | 65 | 65 | 65 | 65 | 65 |
|  |  | Genetic PD | R | 0.054 | 0.138 | -0.045 | 0.367 | 0.074 | 0.049 | -0.033 | 0.088 | -0.273 |
|  |  |  | *p* value | 0.674 | 0.277 | 0.725 | **0.003** | 0.561 | 0.703 | 0.798 | 0.488 | **0.029** |
|  |  |  | N | 64 | 64 | 64 | 64 | 64 | 64 | 64 | 64 | 64 |
|  | Female | Idiopathic PD | R | -0.006 | -0.013 | -0.023 | 0.173 | -0.062 | 0.252 | -0.008 | -0.130 | -0.028 |
|  |  |  | *p* value | 0.967 | 0.925 | 0.866 | 0.202 | 0.650 | 0.061 | 0.953 | 0.339 | 0.838 |
|  |  |  | N | 56 | 56 | 56 | 56 | 56 | 56 | 56 | 56 | 56 |
|  |  | Genetic PD | R | 0.163 | 0.150 | 0.154 | 0.269 | 0.194 | -0.060 | 0.246 | 0.219 | 0.110 |
|  |  |  | *p* value | 0.214 | 0.253 | 0.242 | **0.038** | 0.137 | 0.649 | 0.058 | 0.093 | 0.404 |
|  |  |  | N | 60 | 60 | 60 | 60 | 60 | 60 | 60 | 60 | 60 |

Abbreviations: N, number of subjects; MDS-UPDRS III, Movement Disorders Society Unified Parkinson's Disease Rating Scale, part III. All *p* values from Spearman's correlation are shown. Significant *p*-values from Spearman's correlation that remained significant after partial correlation analysis adjusted for age, disease duration, and LEDD are shown in bold.

No significant correlations were observed in iPD patients **(Figure 7a,b**; **Table 5)**. Conversely, a positive correlation between D-Ser levels and motor symptom severity was observed in gPD patients of both sexes **(Figure 7c,d**; **Table 5)**. Additionally, a negative correlation between the L-Gln/L-Glu ratio and motor symptom scores was observed in gPD males **(Table 5)**.

**Figure 7.**
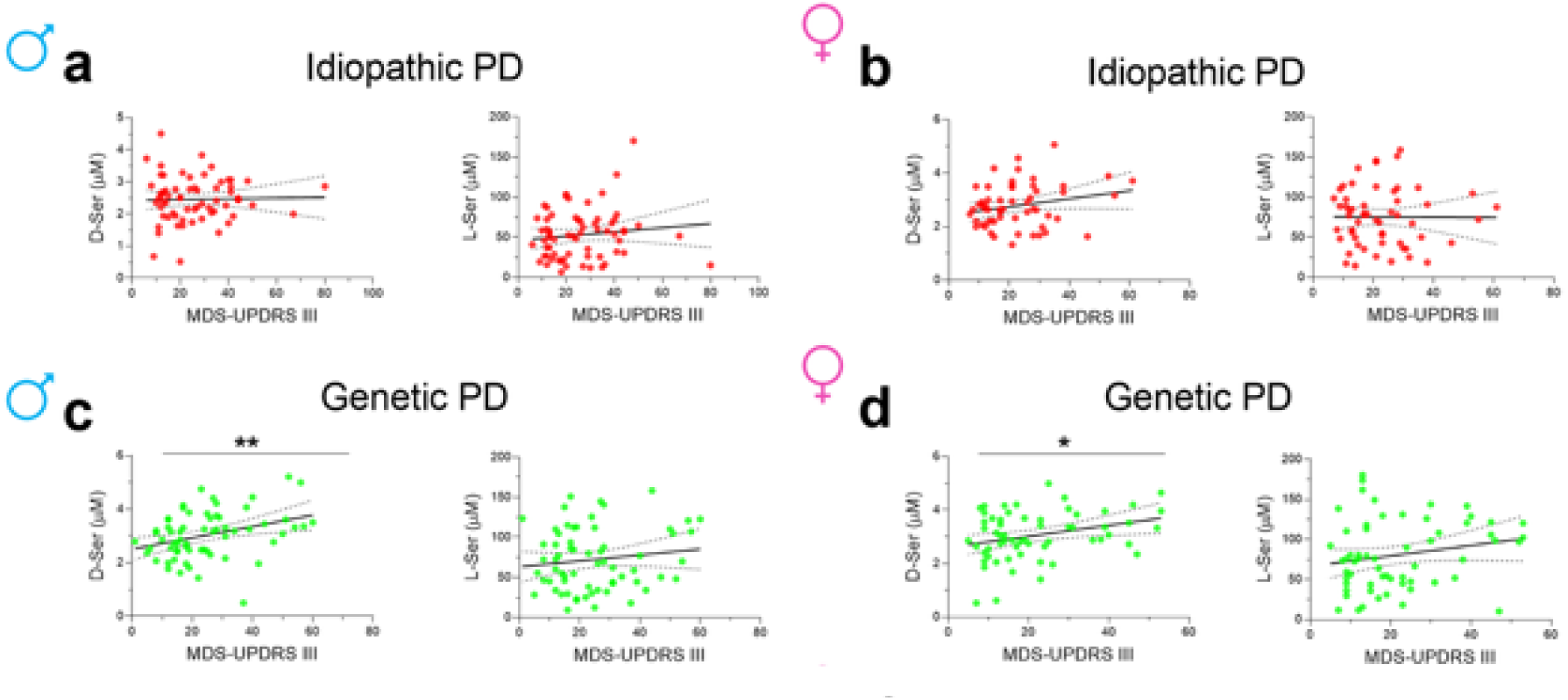
Correlation of serum D-serine and L-serine concentrations with MDS-UPDRS III of idiopathic PD and genetic PD patients stratified for sex. Scatterplots showing the association of D-serine (D-Ser) or L-serine (L-Ser) content with MDS-UPDRS III of male (**a, c, e)** and female (**b, d, f**) idiopathic PD (**a, b**), genetic PD (**c, d**). Best fit lines and 95% confidence intervals are shown. \**p* < 0.05,** *p* < 0.01, Spearman’s correlation test confirmed by partial correlation analysis.

Stratification by genotype revealed that the correlation between D-Ser levels and motor symptom severity was mainly driven by *TMEM175* mutation carriers, irrespective of sex **(Figure S5)**.

These findings suggest that the relationship between circulating NMDAR-related amino acids and motor severity is influenced by genetic background.

### NMDAR subunit gene variants exhibit sex-and subtype-specific association in Parkinson’s disease

Based on the sex-and PD subtype-related effects observed in serum NMDAR-related amino acid levels, we investigated whether these factors also influence variations in NMDAR subunit genes, including *GRIN1*, *GRIN2A*, and *GRIN2B*.

Principal Component Analysis (PCA) confirmed that the study cohort was genetically homogeneous **(Figure S6a)**, with PC1 and PC2 explaining 25% of variance **(Figure S6b)**. All analyses were adjusted for age, sex and 10 principal components.

We then tested rare exonic non-synonymous variants using Optimal Unified Approach for Rare-Variant Association (SKAT-O) in the same cohort used for HPLC analysis, stratified by sex and PD subtype. No significant associations were observed.

Next, we analyzed common variants (Minor Allele Frequency (MAF) > 0.01, referred to our internal database) across *GRIN1*, *GRIN2A*, and *GRIN2B* regions (including exons, UTRs and intronic regions near splice sites; n = 70 variants from GRCh38 Whole Exome Sequencing (WES) data). In the discovery cohort, nominal associations were observed in male iPD patients for *GRIN2A* rs11866570 (p value (p) = 0.007, Odd Ratio (OR) = 0.24) and *GRIN2B* rs11055581 (p = 0.01, OR = 2.15), while no associations were detected in females **(Table 6)**. In gPD patients, sex-specific nominal associations were also observed across both genes (*GRIN2A* rs62621078: p = 0.005, OR = 1.23; *GRIN2B* rs7301328: p = 0.01, OR = 0.46 in males; *GRIN2A* rs6497540: p = 0.004, OR = 0.38; *GRIN2B* rs1806201: p = 0.004, OR = 0.35 in females) (**Table 6**). No variants in *GRIN1* were associated with PD. However, none of these associations survived correction for multiple testing (threshold 7.1 × 10⁻⁴) and should therefore be considered exploratory.

**Table 6.** Common variants in *GRIN2A* and *GRIN2B* genes encoding NMDAR subunits associated with PD in case-control study.

| Sex Cohort (N) | Contrast (N) | Gene | chr | Position hg38 | nt change | position | SNP id | test | p | OR |
| --- | --- | --- | --- | --- | --- | --- | --- | --- | --- | --- |
| Male PD vs HC (129 vs 80 ) | Idiopathic PD vs HC (65 vs 80) | GRIN2A | 16 | 10112318 | T>C | Intronic | rs11866570 | ADD | 0.007 | 0.245 |
|  |  | GRIN2B | 12 | 13675725 | T>C | intronic | rs11055581 | ADD | 0.01 | 2.15 |
|  | genetic PD vs HC (64 vs 80) | GRIN2A | 16 | 10112087 | C>T | Intronic | rs62621078 | ADD | 0.005 | 1.23 |
|  |  | GRIN2B | 12 | 13865843 | G>C | SV p.P122P | rs7301328 | ADD | 0.01 | 0.46 |
| Female PD vs HC (116 vs 123) | Idiopathic PD vs HC (56 vs 123) | none | - | - | - | - | None | ADD | NS | NS |
|  | genetic PD vs HC (60 vs 123) | GRIN2B | 12 | 13564574 | G>A | SV p.T888T | rs1806201 | ADD | 0.004 | 0.35 |
|  |  | GRIN2A | 16 | 9841209 | T>G | Intronic | rs6497540 | ADD | 0.004 | 0.38 |
| Male MNI-PD subtype vs MNI-HC (259 vs 108) | Idiopathic PD vs HC (120 vs 108) | GRIN2A | 16 | 10112318 | T>C | Intronic | rs11866570 | ADD | 0.005 | 0.242 |
|  |  | GRIN2B | 12 | 13675725 | T>C | intronic | rs11055581 | ADD | NS | NS |
|  | genetic PD vs HC (139 vs 108) | GRIN2A | 16 | 10112087 | C>T | Intronic | rs62621078 | ADD | NS | NS |
|  |  | GRIN2B | 12 | 13865843 | G>C | SV p.P122P | rs7301328 | ADD | NS | NS |
| Female MNI-PD subtype vs MNI-HC (161 vs 174) | Idiopathic PD vs HC (73 vs 174) | none | - | - | - | - | None | ADD | NS | NS |
|  | genetic PD vs HC (88 vs 174) | GRIN2B | 12 | 13564574 | G>A | SV p.T888T | rs1806201 | ADD | 0.008 | 0.5 |
|  |  | GRIN2A | 16 | 9841209 | T>G | Intronic | rs6497540 | ADD | NS | NS |

Abbreviations: N, number of subjects; PD, Parkinson's disease; HC, Healthy controls; MNI, Mediterranean Neurological Institute; chr, chromosome; position hg38, genomic position human genome assembly 38; nt change, nucleotide change; SNP id, single nucleotide polymorphism identification number; p, p-value calculated with logistic regression model with Plink software; ADD, additive genetic model; OR, Odds Ratio; SV, synonymous variant; NS, not significant. Significance after Bonferroni correction $p=7.1E-04$ . Genes analysed: *GRIN1*, *GRIN2A*, *GRIN2B*

Validation in the MNI stratified cohort confirmed the association of *GRIN2A* rs11866570 in male iPD patients (p = 0.005, OR = 0.24), and *GRIN2B* rs1806201 in female gPD patients (p = 0.008, OR = 0.5) **(Table 6)**.

In larger, non-stratified cohorts (MNI and PDGC/UK biobank, see Methods for details), one *GRIN2B* variant (rs11055581) showed consistent association across datasets (MNI: p = 0.003, OR = 1.48; PDGC: p < 0.00001, OR = 1.12), while other variants showed weaker or inconsistent effects **(Table S13)**.

Overall, these results suggest a potential but preliminary involvement of *GRIN2B* in PD susceptibility, while *GRIN2A* may act as a context-dependent modifier. All findings should be interpreted cautiously, given the lack of significance after multiple testing correction.

Finally, to explore the functional impact of PD-associated GRIN2A and GRIN2B variants, we queried the GTEx portal (https://www.gtexportal.org/home/) across multiple brain regions. Although GTEx data are not stratified by sex or PD subtype, protective *GRIN2A* (rs11866570) and *GRIN2B* (rs7301328 and rs1806201) variants were generally associated with increased gene expression across basal ganglia and related brain regions, except for reduced *GRIN2A* expression in the caudate (**Figure 8a-c**; **Tables S14-S16)**. These findings suggest a potential link between reduced *GRIN2A/GRIN2B* expression and increased PD risk.

**Figure 8.**
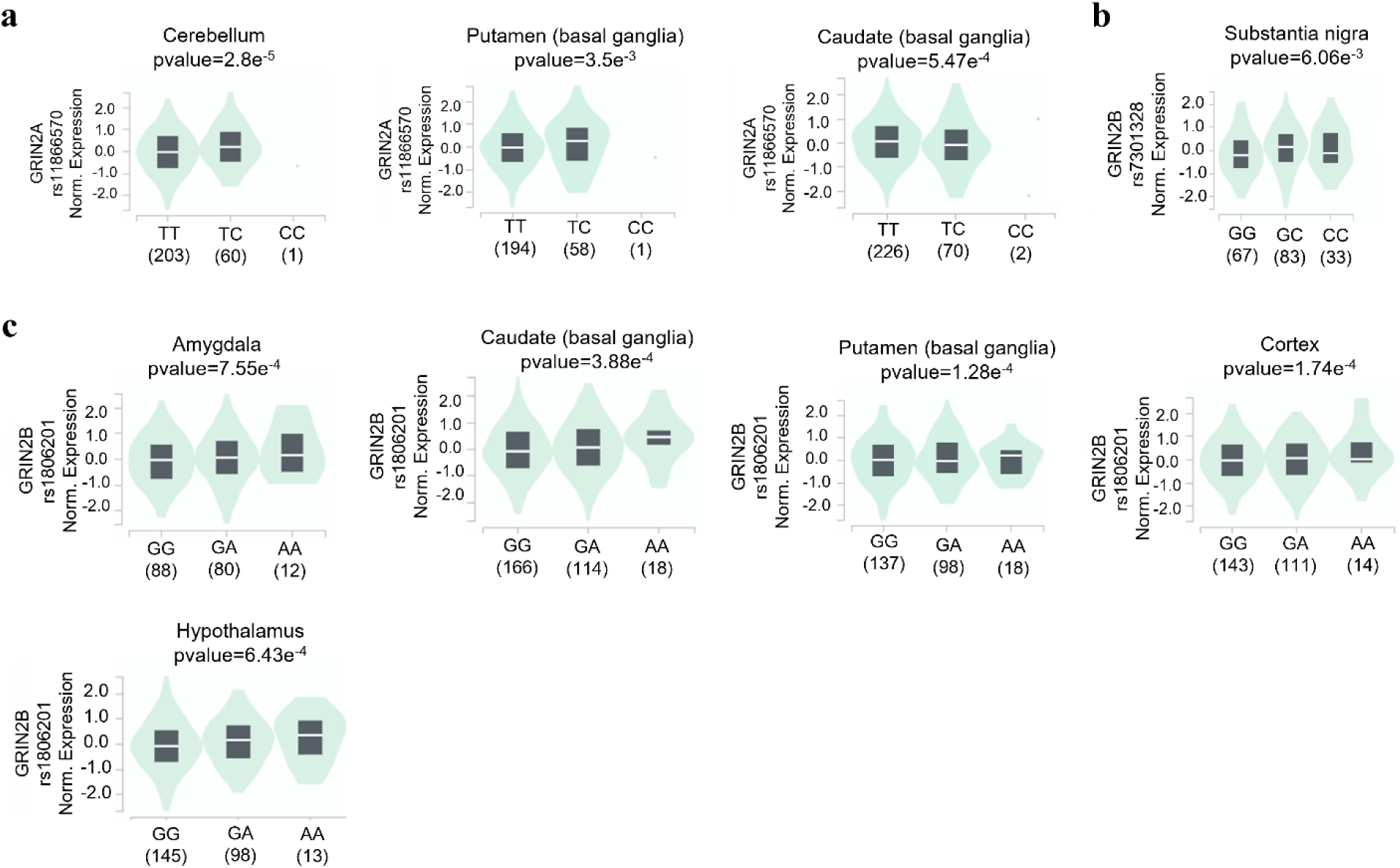
Genotype-Tissue Expression (Gtex) analysis of GRIN2A and GRIN2B transcripts in human brain regions associated with the presence of the allele variants rs11866570-C, rs7301328-C and rs1806201-A. Violin plots show significantly increased expression of *GRIN2A* transcript associated with the presence of the protective allele C in Cerebellum and Putamen **(a)**. On the contrary, reduced expression was observed in Caudate **(a)**. In **b** is shown the significant increased expression of *GRIN2B* transcript in the Substantia nigra in the presence of the protective allele rs7301328-C. In **c** is reported the increased expression of *GRIN2B* transcript in Amygdala, Caudate, Putamen, Cortex and Hypothalamus associated with the protective allele rs1806201-A.

## Discussion

Previous studies have reported significant alterations in blood amino acid levels in PD patients^33,34,36–38,42^, although findings have been inconsistent across cohorts and analytical platforms^38,39^. In particular, the contribution of sex and genetic factors to serum NMDAR-related amino acid changes remains poorly defined.

In the present study, we addressed this gap by combining HPLC-based quantification with genetic and clinical data from a large cohort of PD patients and a balanced sex-matched group of healthy controls.

Our analyses identify a previously underappreciated sex-based pattern of NMDAR-related amino acid dysregulation in PD. Alterations were largely driven by male patients and were most pronounced in idiopathic PD, whereas female patients and genetic forms showed limited or no significant changes. Specifically, male idiopathic PD patients showed marked reductions of D-Ser, L-Ser, Gly, L-Glu, L-Gln, L-Asp and L-Asn, together with an increased D-Ser/total Ser ratio, compared to sex-matched healthy controls.

Integrated modelling further confirmed that sex represents the primary determinant of circulating NMDAR-related amino acid variation in PD, while disease subtype contributes to a lesser extent. Importantly, the absence of a significant subtype × sex interaction indicates that the pronounced alterations observed in idiopathic male patients reflect additive effects of sex and disease subtype, rather than a true interaction between these variables.

The marked downregulation of NMDAR-related amino acid content observed in male idiopathic PD patients is consistent with previous metabolomic and HPLC studies reporting sex-specific differences in amino acid and lipid metabolism in PD^35,39,41,42,44–46^. While hormonal, genetic, epigenetic, and nutritional factors may contribute to this sexual dimorphism, further studies are required to identify the underlying mechanisms that confer protection in females against the NMDAR-related disruption of amino acid homeostasis reported in male patients.

In contrast, our HPLC analyses showed comparable serum NMDAR-related amino acid levels in patients carrying at least one mutation in *LRRK2*, *TMEM175*, *GBA1*, or *PARK2/PINK1/PARK7*, compared with healthy controls. Although these genes are widely expressed in both the CNS and peripheral organs (https://www.proteinatlas.org/)^47–55^, and implicated in mitochondrial function, lysosomal activity, autophagy, and oxidative stress regulation^53,56–61^, our findings suggest that, individually, they do not produce detectable changes in circulating NMDAR-related amino acid levels under HPLC measurement conditions.

Taken together, our results suggest that systemic alterations in NMDAR-related amino acid homeostasis may emerge from the combined effect of multiple risk factors, including genetic, sex and environmental influences, as observed in idiopathic male patients. This integrated perspective may help clarify inconsistencies across previous blood-based studies^38,39^ lacked sex and subtype stratification.

Increasing evidence supports the view of PD as a complex, multisystem disorder ^29,30^. In this context, circulating amino acids are closely linked to systemic metabolic homeostasis, peripheral organ function, diet, and gut microbiota composition^62–68^. Accordingly, the broad deficiency of NMDAR-related amino acids observed in idiopathic male patients, also relative to the genetic PD group, may reflect a generalized metabolic dysregulation and represent a potential blood-based signature for patient stratification.

Beyond their role in neurotransmission, the reductions in L-Glu, L-Gln, L-Asp and Gly observed in idiopathic male patients suggest a perturbation of metabolic pathways involved in energy balance and redox homeostasis. Indeed, L-Glu and Gly are key components of glutathione synthesis, and their reduction may reflect impaired antioxidant capacity. Supporting this interpretation, previous analyses revealed alterations in glutathione metabolism in PD^69,70^. In addition, L-Glu and L-Asp play a central role in mitochondrial metabolism, supporting the hypothesis of a broader metabolic imbalance in PD^36,37,41,71–74^.

The combined reduction of L-Ser, D-Ser and Gly in idiopathic male patients further supports a perturbation of serine-glycine metabolism, a pathway implicated in multiple cellular processes, including membrane phospholipids and myelin biosynthesis, one-carbon metabolism and neuroinflammation^75–78^. Notably, our findings in the serum of male iPD patients contrast with previous reports of increased serine enantiomer levels in the caudate-putamen and cerebrospinal fluid ^18–21^, suggesting that reduced circulating serine levels primarily reflect systemic metabolic dysfunction rather than direct basal ganglia imbalance.

Our results may have potential clinical implications. Reduced serum levels of NMDAR-related amino acids or their precursors in idiopathic male patients may support patient stratification and help identify individuals who could benefit from targeted amino acid interventions^79–80^. In this context, modulation of NMDAR co-agonists has been explored as a therapeutic strategy. Oral D-Ser supplementation, for instance, has shown beneficial effects in a small PD cohort, improving both motor and non-motor symptoms as an adjunct to standard therapy^81,82^. However, these findings remain preliminary and require validation in larger and longitudinal clinical studies.

We also observed a positive correlation between age and D-Ser levels, as well as D-Ser/total Ser ratio, in female healthy controls and idiopathic patients, consistent with a previous report ^37^, suggesting a physiological sex-dependent effect. In addition, the correlation between D-Ser levels and motor symptom severity in genetic PD, particularly in *TMEM175* mutation carriers, may indicate a link between amino acid regulation and disease progression in specific subgroups. However, given the cross-sectional design and limited sample size of our analysis, these findings require further validation.

Finally, our genetic analyses indicate that *GRIN2B* variants showed nominal associations with PD across cohorts, suggesting a potential role in disease susceptibility. However, these associations did not survive correction for multiple testing and should therefore be considered exploratory. Consistently, GTEx data (not stratified by sex or PD subtype) indicate that alleles associated with lower PD risk tend to be linked to higher *GRIN2B* expression in relevant brain regions, although this remains correlative. By contrast, *GRIN2A* variants were not associated in non-stratified analyses and instead showed subtype-and sex-dependent trends, supporting a context-dependent modifier role. Overall, these findings suggest a tentative link between genetic and metabolic alterations in the NMDAR pathway, which requires validation in larger, stratified cohorts.

A major strength of our study is its integrated design, combining targeted amino acid quantification, high-resolution genotyping^83,84^ and detailed demographic and clinical phenotyping in a large and well-characterized PD cohort. This approach allowed the identification of sex-and subtype-specific signatures impacting NMDAR-related amino acid metabolism. However, some limitations must be acknowledged. The cross-sectional design precludes causal inference, the absence of drug-naïve patients limits generalizability, and the inclusion of only Caucasian individuals restricts applicability to other populations.

In conclusion, our findings highlight the importance of considering sex and genetic variation in PD research and advocate for the integration of metabolic data to identify reliable biomarkers and therapeutic targets.

## Methods

### Study participants

#### PD-cohort

This is a case-control observational study. 245 independent and unrelated PD patients (129 males and 116 females; 86 familial and 159 sporadic cases) were enrolled in this study. These patients were part of the PD biobank of IRCCS Neuromed/IGB-CNR. Only individuals aged ≥40 years were included. The median age at diagnosis was 62.0 years (IQR 55.0-66.0). All the subjects were of European ancestry and were evaluated by qualified neurologists of the Parkinson Centre of the IRCCS INM Neuromed from June 2015 to December 2017, and from June 2021 to December 2023, with a thorough protocol comprising neurological examination and evaluation of non-motor domains. Information about family history, demographic characteristics, anamnesis, and pharmacological therapy was also collected (the treatment of the PD groups consisted for the most part of a combination of levodopa and dopamine agonists)^83^. Clinical criteria for diagnosis required the presence of at least two cardinal motor signs: asymmetric resting tremor, bradykinesia and rigidity, as well as a good response to levodopa and absence of other atypical features and causes of parkinsonism. Exclusion Criteria for enrolment were: *i*) pre-existing psychiatric conditions; *ii*) presence of neurodegenerative neurological diseases such as multiple sclerosis, lateral sclerosis amyotrophic, Alzheimer’s, neuromuscular pathologies, epilepsy; *iii*) diagnosis of dementia; *iv*) depression; *v*) prolonged intake of anxiolytics, antidepressants, antipsychotics, hypnotic drugs, cognitive stimulants.

The Movement Disorder Society revised version of the Unified Parkinson’s Disease Rating Scale Part III (33 items, maximum score 132; hereafter called UPDRS)^85^ was used to assess clinical motor symptoms. These included language, facial expressions, tremor, rigidity, agility in movements, stability, gait and bradykinesia. Patients were analyzed during the ON period.

The PD cohort includes 121 idiopathic and 124 genetic PD patients, carrying at least one pathogenic mutation in the most frequently mutated genes *LRRK2*, *GBA1*, *TMEM175* and *PARK2/PINK1/PARK7* (see next paragraph “Study Cohort and stratification factors” for the criteria used for patients’ selection). This cohort was used both for quantification of NMDAR-related amino acids by HPLC analysis and for genetic association analysis of *GRIN1*, *GRIN2A* and *GRIN2B* genes.

#### Ethical Compliance

All procedures involving human participants were approved by the Institutional Review Board of the IRCCS Neuromed Italy. The study protocols N°9/2015, N°19/2020, N°4/2023 have been registered in clinicaltrial.gov with the numbers NCT02403765 (Release Date: 04/01/2015), NCT04620980 (Release Date: 11/03/2020), NCT05721911 (Release Date: 30/01/2023).

Clinical investigations were conducted according to the principles expressed in the Declaration of Helsinki. Written informed consent was obtained from all participants.

The research was carried out following the recommendations set out in the Global Code of Conduct for Research in Resource-Poor Settings.

#### Reporting Standards

“Reporting adheres to the STROBE guidelines for case-control studies and the MSI guidelines for biochemical data.”

No formal sample size calculation was performed; the sample size was determined based on the availability of well-characterized subjects from the biobank.

#### HCs cohort

203 neurological controls (80 males and 123 females; median age 61.0 years; IQR 55.0-68.0) were selected for this study. HCs matched for sex and age with PD patients and were negative for mutation/variant in PD genes (see next paragraph “Study Cohort and stratification factors” for the criteria used for patients’ selection). This HCs cohort was used for both HPLC analysis and for genetic association analysis of NMDAR genes.

To minimize potential sources of bias, standardized protocols for clinical assessment, sample collection, and processing were applied. Laboratory analyses were performed on anonymized samples. Sex-matched controls were included to reduce confounding effects.

### Study Cohort and stratification factors

Here, we classified patients based on their genetic background as “idiopathic-PD” and “genetic PD”. We choose not to use the classification “familial PD” or “sporadic PD” since this classification does not always reflect the presence/absence of mutation/variant. Indeed, in our cohort a consistent number of patients (41.1 %) mutated in *LRRK2* were classified as “sporadic” at interview, and similar percentages (ranging from 50 to 65 %) emerged for patients carrying single pathogenic mutation in *GBA1, PARK2, PINK1, PARK7*, and *TMEM175*. This could depend on several factors, such as late age at the onset of PD, the presence of small nuclear families, and the reduced penetrance of PD mutations.

Whole Exome Sequencing (WES) data were used for a comprehensive identification of the genetic background of the study cohort, including PD patients and HCs. Considering the extreme genetic heterogeneity of PD, thirty-seven PD candidate genes were extracted from WES data. The selected genes include those reported in the literature as Mendelian PD genes (*PARK7*/*DJ-1, DNAJC13, DNAJC6, EIF4G1, FBXO7, LRRK2, PARK2, PINK1, SNCA*) and those reported as PD genes/at risk factors (*AIMP2, ANKK1, ANKRD50, CHMP1A, GBA1, GIGYF2, GIPC1, GRK5, HMOX2, HSPA8, HTRA2, IMMT, KIF21B, KIF24, MAN2C1, PACSIN1, RHOT2, SLC25A39, SLC6A3, SLC6A3, SNCAIP, SPTBN1, TMEM175, TOMM22, TVP23A, UCHL1, VPS8, ZSCAN21*)^51,83,86–93^.

The cohort (both PD patients and HCs) was analyzed for the presence of mutations (as reported in the literature or public mutation databases) or rare (Minor allele frequency (MAF) < 0.001, based on gnomAD v.4.1.0) and highly deleterious exonic variants (including non-synonymous, splicing and indels; Combined Annotation Dependent Depletion (CADD) score ≥15) in the panel of the selected PD genes.

We selected 121 PD patients that we named “idiopathic”, in which no mutation/variant was identified in the selected PD genes.

The second group, that we named “genetic PD”, includes 124 PD-affected subjects carrying of at least 1 pathogenic mutation in one of the most frequently mutated PD genes, which includes 17 patients mutated in *LRRK2*, 30 mutated in *GBA1*, 40 with at least 1 mutation in *PARK2/PINK1/PARK7*, 33 mutated in *TMEM175,* and four patients carrying two mutations in PD genes such as *GBA1/TMEM175*, *GBA1/PARK2*, *GBA1/PINK1* and *TMEM175/PARK2*, respectively. No pathogenic mutation was identified in *SNCA*, *VPS35*, *DNAJC13, DNAJC6, EIF4G1, FBXO7* and *UCHL1*, in our cohort, by surveying the public mutations databases (LOVD - An Open Source DNA variation database system (https://lovd.nl/) (**Table S1**).

In particular, the seventeen *LRRK2* mutated patients were carriers of the p.G2019S (14 patients), p.R1441C (2 patients) and p.A419V (1 patient) mutations. Thirty-three *GBA1* mutated patients were carriers of the following pathogenic mutations: p.N409S (9 patients), p.E365K (7 patients), p.T408M (3 patients), p.L483P (2 patients), p.H294Q (2 patients) and p.R17G, p.D66E, p.V230E, p.P240S, p.H367Y, p.S403T, p.E427K, p.D448H, p.V499L, p.K505N (1 patient each). Forty-two PD patients were carriers of at least one pathogenic mutation in one of the recessive PD genes *PARK2*, *PINK1*, *PARK7.* 18 out of 42 PD patients were mutated in *PARK2* including one homozygous patient (p.W453X/ p.W453X), 1 compound heterozygous (p.R275W/exon deletion) and 16 carrying a single heterozygous mutation such as p.R402C (4 patients), p.A82E (5 patients), and p.M192L, p.R234Q, p.T240M, p.V244L, p.R275W, p.E409X, p.T415N, p.W453X (1 patient each). Nine patients were carriers of a single heterozygous mutation in *PINK1* including p.P196L (2 patients), p.W437X (2 patients), p.A168P, p.K186N, p.A291D, p.R326C, p.D525N (1 patient each). Sixteen patients were carriers of a single heterozygous variant in *PARK7* of which 14 carrying the p.R98Q variant and 2 carrying the splicing variant c.252+2->A. Thirty-five PD patients were carriers of one pathogenic mutation in *TMEM175* (as reported in our previous study^51^ which includes p.V147Dfs*104 (4 patients), p.A429Qfs*120 (3 patients), p.A149Gfs*97 (3 patients), p.R35C (3 patients), p.L405V (4 patients), p.R414W (3 patients), p.R335H (3 patients), p.T105A (2 patients), p.I78T (2 patients), and p.R260C, p.A270T, p.I280M, p.P286L, p.A326V, p.S348L, p.A424V, p.R481W (1 patient each) (Table S1).

The HCs that we included in the study were negative for mutation/variant in the selected panel of PD genes.

### Collection and storage of serum samples

Blood sampling was performed after a 6-h fasting. Whole blood was collected by peripheral venipuncture into clot activator tubes and gently mixed.

Sample was stored upright for 30 min at room temperature to allow blood to clot and centrifuged at 2000 ×g for 10 min at room temperature. Serum was aliquoted (0.5 ml) in polypropylene cryotubes and stored at −80 °C before usage. Unique anonymized codes have been assigned to the samples for processing and subsequent analysis, maintaining the confidentiality of personal data.

### HPLC analysis of amino acids content

Serum samples (100 μl) were mixed in a 1:10 dilution with HPLC-grade methanol (900 μl) and centrifuged at 13,000 ×g for 10 min; supernatants were dried and then suspended in 0.2 M trichloroacetic acid (TCA). TCA supernatants were then neutralized with 0.2 M NaOH and subjected to precolumn derivatization with o-phthaldialdehyde /N-acetyl-L-cysteine in 50% methanol. Amino acids derivatives were resolved on a UHPLC Nexera X3 system (Shimadzu) by using a Shimpack GIST C18 3-μm reversed-phase column (Shimadzu, 4.0 × 150 mm) under isocratic conditions (0.1 M sodium acetate buffer, pH 6.2, 1% tetrahydrofuran, and 1 ml/min flow rate). A washing step in 0.1 M sodium acetate buffer, 3% tetrahydrofuran and 47% acetonitrile, was performed after every run. Identification and quantification of amino acids were based on retention times and peak areas, compared with those associated with external standards. The detected amino acids concentration was expressed as μM.

### Statistical analysis

The primary outcomes were the amino acid concentrations. The main exposures were PD subtype (iPD and gPD) and sex. Covariates included age, disease duration, and L-DOPA equivalent daily dose (LEDD).

Clinical and demographic characteristics were described as median and the interquartile range (IQR) or absolute frequencies. Comparisons between PD patients and HCs were evaluated using the Mann-Whitney U or Kruskal-Wallis test for continuous variables and Chi-Square test for dichotomous variables. Comparison of serum amino acid levels between PD and HCs groups was first performed using a Mann-Whitney U or Kruskal-Wallis test, followed by Dunn’s multiple comparisons post hoc, when required. An ANCOVA model with “diagnosis” as a between-factor and age, sex and LEDD as covariates was used to control for the effect of these factors on serum amino acid levels. For sex stratified analysis, age-and LEDD-adjusted ANCOVA model was used.

The correlation of serum amino acid concentration with age, age at PD onset, disease duration, LEDD and MDS-UPDRS III was evaluated with Spearman’s correlation test. Partial correlation analyses adjusted for the effect of potential confounders (age, disease duration, LEDD) were adopted to confirm the correlation between serum amino acid levels and PD clinical features (disease duration, LEDD and MDS-UPDRS III). Significance was set at p < 0.05 for all analyses. Data were analysed by using SPSS 26.0 software (IBM, Armonk, NY, USA).

Two sets of linear models were fitted to evaluate the effects of sex and disease subtype on serum NMDAR-related amino acid levels. Amino acid concentrations were natural log-transformed prior to analysis. All models were estimated using ordinary least squares with heteroscedasticity-consistent (HC3) robust standard errors. In the full cohort (PD + HC), Model 1 included group (PD vs HC), sex, age, and a group × sex interaction term. The interaction coefficient was used to assess whether the PD–HC difference differed by sex. In PD patients only, Model 2 included subtype (iPD vs gPD), sex, age, disease duration, LEDD, and a subtype × sex interaction term. To allow sex-specific interpretation of subtype effects, models were fitted using alternative reference categories for sex. Collinearity among covariates was assessed, and highly correlated variables were excluded. P-values were adjusted for multiple testing across outcomes using the Benjamini–Hochberg false discovery rate (FDR) procedure. Statistical significance was defined as p_FDR < 0.05.

### Study cohorts used for genetic analysis

#### MNI-PD cohort

The Mediterranean Neurological Institute (MNI)-PD cohort included 804 independent and unrelated PD patients (501 males; 300 familiar and 504 sporadic cases), for which Whole exome sequencing (WES) data are available. This cohort is part of the Parkinson’s disease Biobank of the IRCCS Neuromed and of the Institute of Genetics and Biophysics (CNR).

All the subjects were of European ancestry and were evaluated by qualified neurologists of the Parkinson Centre of the IRCCS INM Neuromed from June 2015 to December 2017, and from June 2021 to December 2023, with a thorough protocol comprising neurological examination and evaluation of non-motor domains. Information about family history, demographic characteristics, anamnesis, and pharmacological therapy was also collected (the treatment of the PD groups consisted for the most part of a combination of levodopa and dopamine agonists)^83^. This cohort was used to identify susceptibility or protective factors for PD.

#### MNI-PD stratified cohort

This cohort was selected from the entire MNI-PD cohort of 804 PD patients with the same criteria used to select the PD patients analyzed by HPLC and includes 193 idiopathic and 227 genetic PD patients carrying at least one pathogenic mutation in the most frequently mutated genes *LRRK2*, *GBA1*, *TMEM175* and *PARK2/PINK1/PARK7* (see the paragraph “Study Cohort and stratification factors” for the criteria used for patient selection). This cohort was used as validation cohort in genetic association analysis to confirm the association with PD in the cohort stratified by gender and genetic background.

#### MNI-HC cohort

282 neurological HCs were recruited by the same group of neurologists, among the patients’ wives/husbands, after having ascertained the lack of neurological pathologies and the absence of affected family members. WES data were available for the entire cohort. The MNI-HC cohort was used for association analysis.

#### Replication cohort

4586 PD patients (from PDGC cohort) and 43989 CNT (from UK biobank) whose data were downloaded from the PDGC Variant browser (https://pdgenetics.shinyapps.io/VariantBrowser/).

### Associations analysis

Principal Component Analysis (PCA) performed with Plink software was used to characterize the genetic diversity of the study sample (PD_MNI, CNT_MNI) (**Figure S6a**)^94^. The analysis was carried out by using common variants (Minor allele frequency MAF > 0.01), PC1 and PC2 were found to contribute to a variance of 25% among samples **(Figure** S**6b**).

To investigate the genetic contribution of rare variants MAF was set to: MAF < 0.01, MAF<0.005 and MAF< 0.001 on the phenotype, gene-based analyses were carried out using the unified Optimal Sequence Kernel Association Test (SKAT-O) of the R SKAT package (http://cran.nexr.com/web/packages/SKAT/index.html), age was used as covariate.

To identify the genetic contribution given by common variants (MAF > 0.01) we adopted a logistic regression model trough plink2 software, by adjusting for age and the 10 principal components; we also adjusted for sex when we analyzed the entire cohort regardless of gender. p-values were adjusted for Bonferroni multiple testing correction.

### Expression studies in human brain regions

The Genotype-Tissue Expression (GTEx) portal (https://www.gtexportal.org/home) was used to obtain gene expression data for the identified SNPs. The analysis included all available adult human brain regions, specifically: amygdala, anterior cingulate cortex (BA24), caudate nucleus, putamen, substantia nigra, cerebellar hemispheres, cerebellum, cerebral cortex, frontal cortex (BA9), hippocampus, hypothalamus, and nucleus accumbens.

## Declaration statements

### Data Availability

The datasets generated and analyzed during the current study are not publicly available due to institutional data management policies and the absence of a dedicated public repository for the specific metabolite quantification datasets generated in this study, but are available from the corresponding author on reasonable request.

### Code Availability

Not applicable

## Data Availability

All data produced in the present work are contained in the manuscript

## Acknowledgments

The authors are grateful to all the patients, their caregivers, the Clinical Parkinson’s Disease Center of IRCCS Pozzilli and the PD biobank of IRCCS Neuromed and IGB-CNR for the kind cooperation with this study. The authors are grateful to Enza Maria Valente and Alberto Imarisio for their valuable discussion.

This study was partially funded by Italian Ministry of University and Research (PRIN 2022 - COD. 2022XF7YYL to AU and PRIN 2022 – COD. 2022W3RKLJ to TE). The work of A.U., T.N. and T.E. was supported by NEXTGENERATIONEU (NGEU) and funded by the Ministry of University and Research (MUR), National Recovery and Resilience Plan (NRRP), project MNESYS (PE0000006) – A Multiscale integrated approach to the study of the nervous system in health and disease (DN. 1553 11.10.2022). The work of T.E. was supported by Next Generation EU - PNRR M6C2 Investimento 2.1 valorizzazione e potenziamento della ricerca biomedica del SSN grant n. PNRR-MAD-2022-12375960 and grant n. PNRR-MCNT2-2023-12377375. TE was also supported by Ministry of Health, Ricerca Corrente. The study of TE was partially funded by Ministry of Enterprises and Made in Italy (MIMIT) project Neurotechno n. F/180029/01/X43.

Funders had no role in the study design, data collection, data analysis, manuscript preparation, or decision to publish.

## Author contributions

IY: Investigation, review & editing; FC: data analysis, review & editing; TN: data analysis, review & editing; MV: data analysis, review & editing; ADM: Investigation, review & editing; NM: review & editing, SP: review & editing; FE: Writing – review & editing; TE: Funding acquisition, Project administration, Supervision, Writing – review & editing. AU: Conceptualization, Funding acquisition, Project administration, Supervision, Writing – review & editing.

