## Supplementary data for "Sex and genotype influence the disruption of circulating NMDAR-related amino acids in patients with Parkinson’s disease"

[4] Clinical Pathology Unit, Fondazione IRCCS Ca' Granda Ospedale Maggiore Policlinico, Milano, Italy.

[5] IRCCS INM Neuromed, 86077, Pozzilli, Italy;

[6] Department of Human Neuroscience, Sapienza University of Rome, Italy;

[7] Department of Agricultural Sciences, University of Naples "Federico II", 80055, Portici, Italy.

*contributed equally and share joint first authorship

[@] to whom correspondence should be addressed

Corresponding authors:

Dr Teresa Esposito PhD Molecular Genetics and Genomics Laboratory Institute of Genetics and Biophysics, "Adriano Buzzati Traverso", Italian National Research Council (CNR) 80131 Naples – Italy. PI IGB-CNR laboratory IRCCS INM Neuromed 86077 Pozzilli – Italy.

Prof Alessandro Usiello PhD. Department of Environmental, Biological and Pharmaceutical Sciences and Technologies (DISTABIF). Università della Campania, L. Vanvitelli, Viale Abramo Lincoln 5, 81100, Caserta, Italy. PI Neuroscience Laboratory. CEINGE, Biotecnologie Avanzate Franco Salvatore, S.c.a.rl. 80145 Naples- Italy

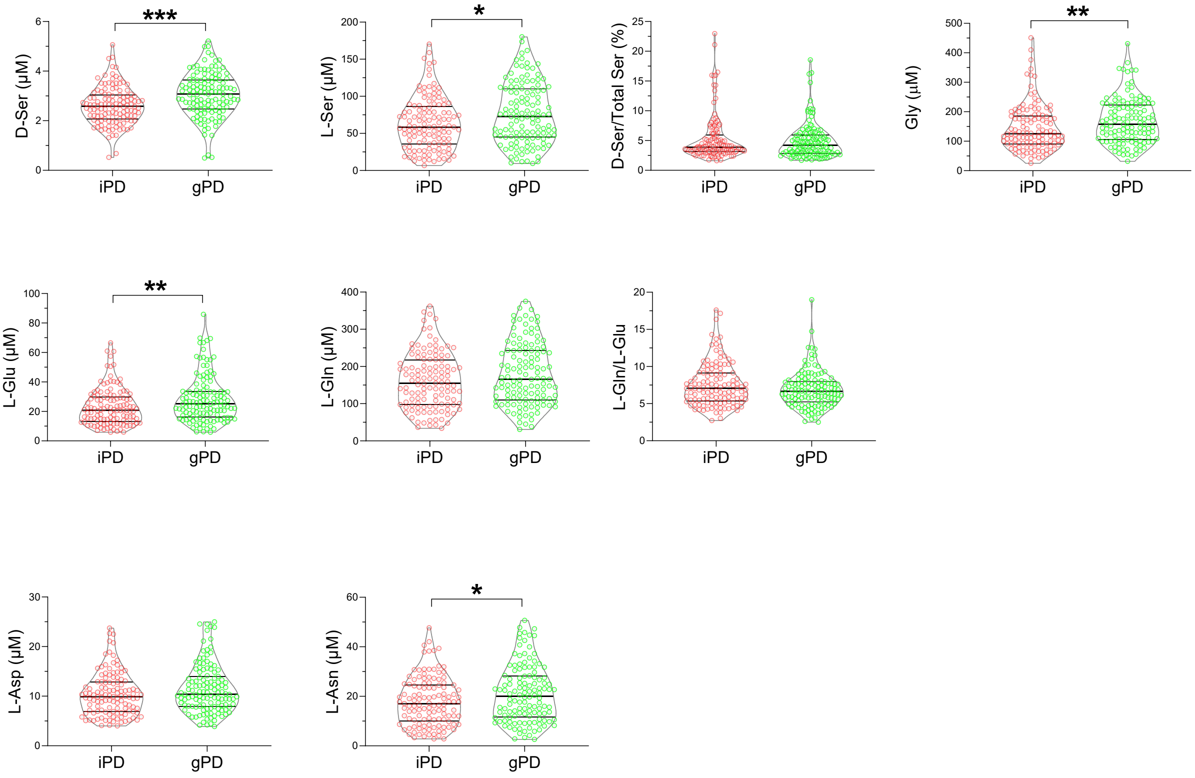


**Figure S1. D- and L- amino acids levels compared between idiopathic and genetic PD.** Analysis of D-serine (D-Ser), L-serine (L-Ser), D-Ser/total Ser ratio, glycine (Gly), L-glutamate (L-Glu), L-glutamine (L-Gln), L-Gln/L-Glu ratio, L-aspartate (L-Asp), and L-asparagine (L-Asn) levels in the serum of idiopathic PD (iPD) and genetic PD patients (gPD). The amino acid content was expressed as µM. In each sample, free amino acids were detected in a single run. Dots represent the single subjects' values, while lines illustrate the median with interquartile range. *$p < 0.05$, *$p < 0.01$, ***$p < 0.001$; Mann-Whitney test following a significant ANCOVA adjusted for age, sex, LEDD, and disease duration.

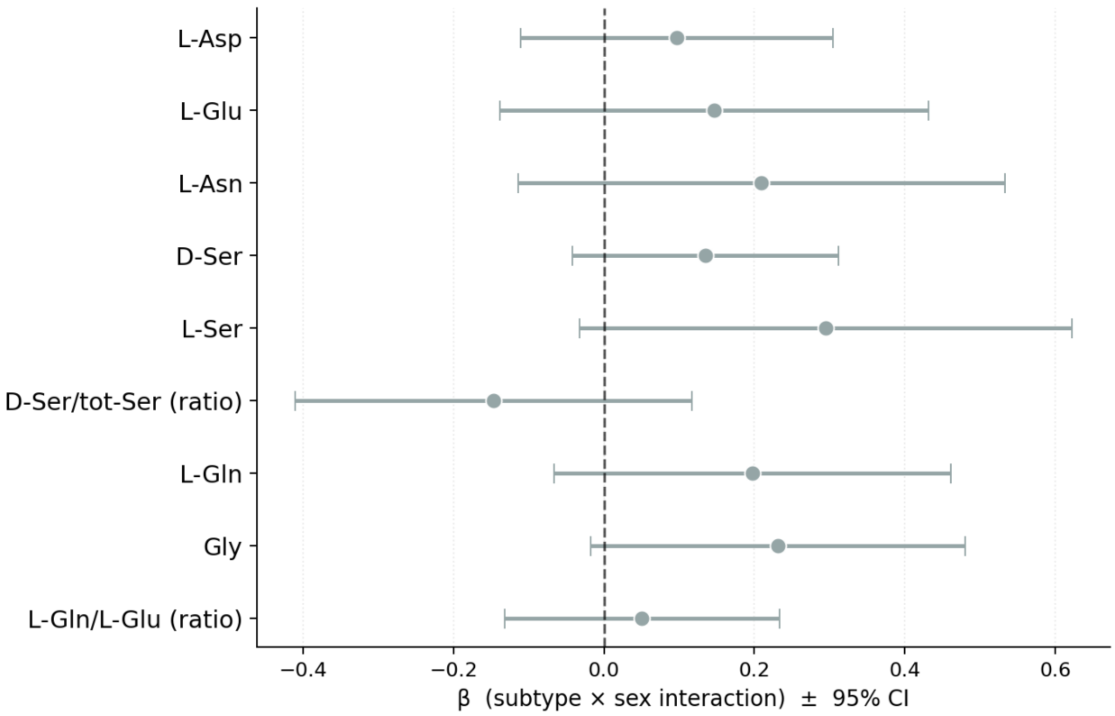


**Figure S2**. **Formal test of subtype × sex interaction on serum NMDAR-related amino acid levels in PD patients.** Forest plot showing the β coefficients of the subtype × sex interaction term from linear models fitted separately for each amino acid outcome: log(amino acid) ~ subtype + sex + age + disease duration + LEDD + subtype:sex, where subtype = iPD (0) vs gPD (1) and sex = female (0) vs male (1). A positive β indicates that the difference in serum levels between gPD and iPD is greater in males than in females. Error bars represent 95% confidence intervals based on heteroscedasticity-consistent (HC3) robust standard errors. No interaction term reached significance after Benjamini-Hochberg false discovery rate (FDR) correction for multiple comparisons (9 tests). Grey = not significant after FDR correction. Analysis restricted to PD patients (n = 245)

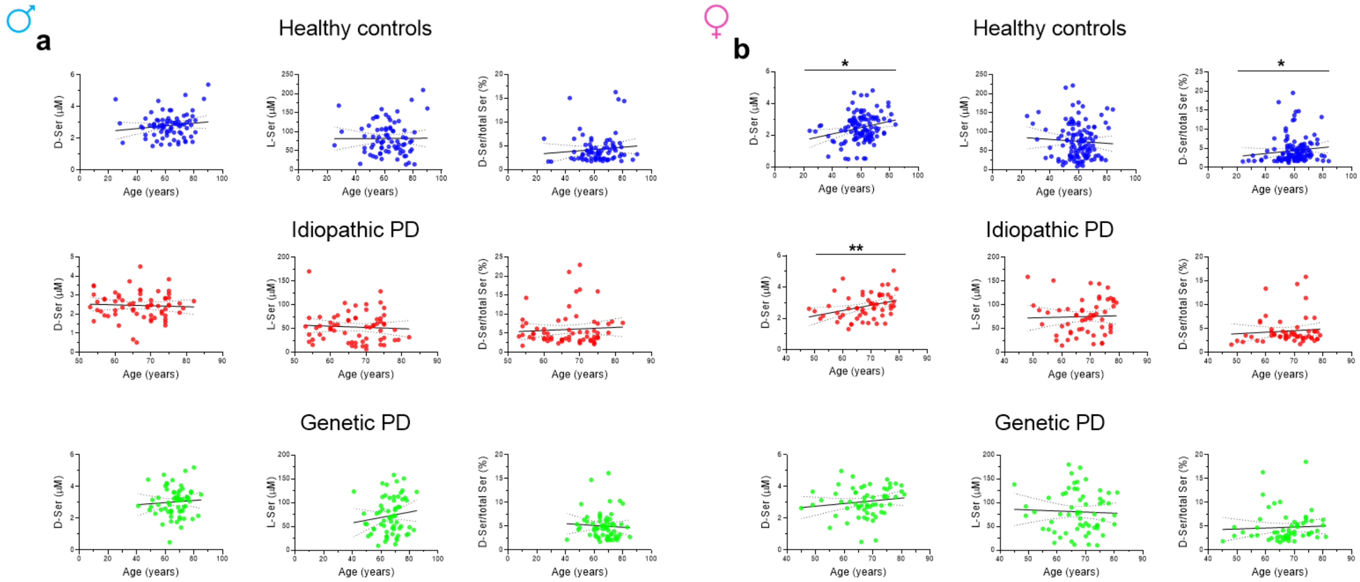


**Figure S3. Correlations between serum D- and L-Ser levels with the age of male and female healthy controls and idiopathic PD patients.** Correlation analysis of D-serine (D-Ser), L-serine (L-Ser), and the D-Ser/total Ser ratio with age was performed in male **(a)** and female **(b)** healthy subjects, idiopathic PD and genetic PD. Best fit lines and 95% confidence intervals are shown. *p<0.05, **p<0.01, Spearman's correlation test.

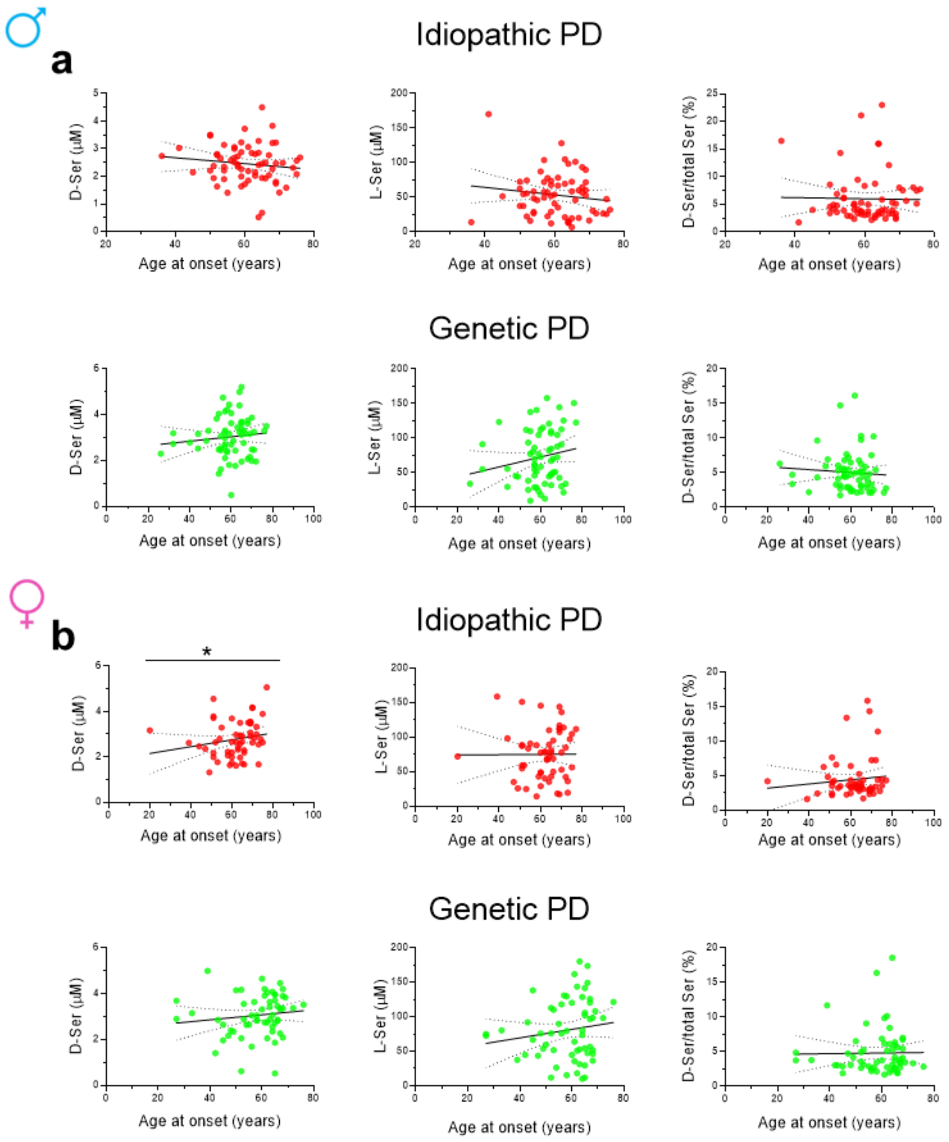


**Figure S4. Association of D-Ser and L-Ser levels with the age at onset within male and female idiopathic PD and genetic PD.** Correlation analysis of D-serine (D-Ser), L-serine (L-Ser), and the D-Ser/total Ser ratio with age at onset in both male **(a)** and female **(b)** idiopathic PD and genetic PD patients. Best fit lines and 95% confidence intervals are shown. *$p<0.05$, Spearman's correlation test.

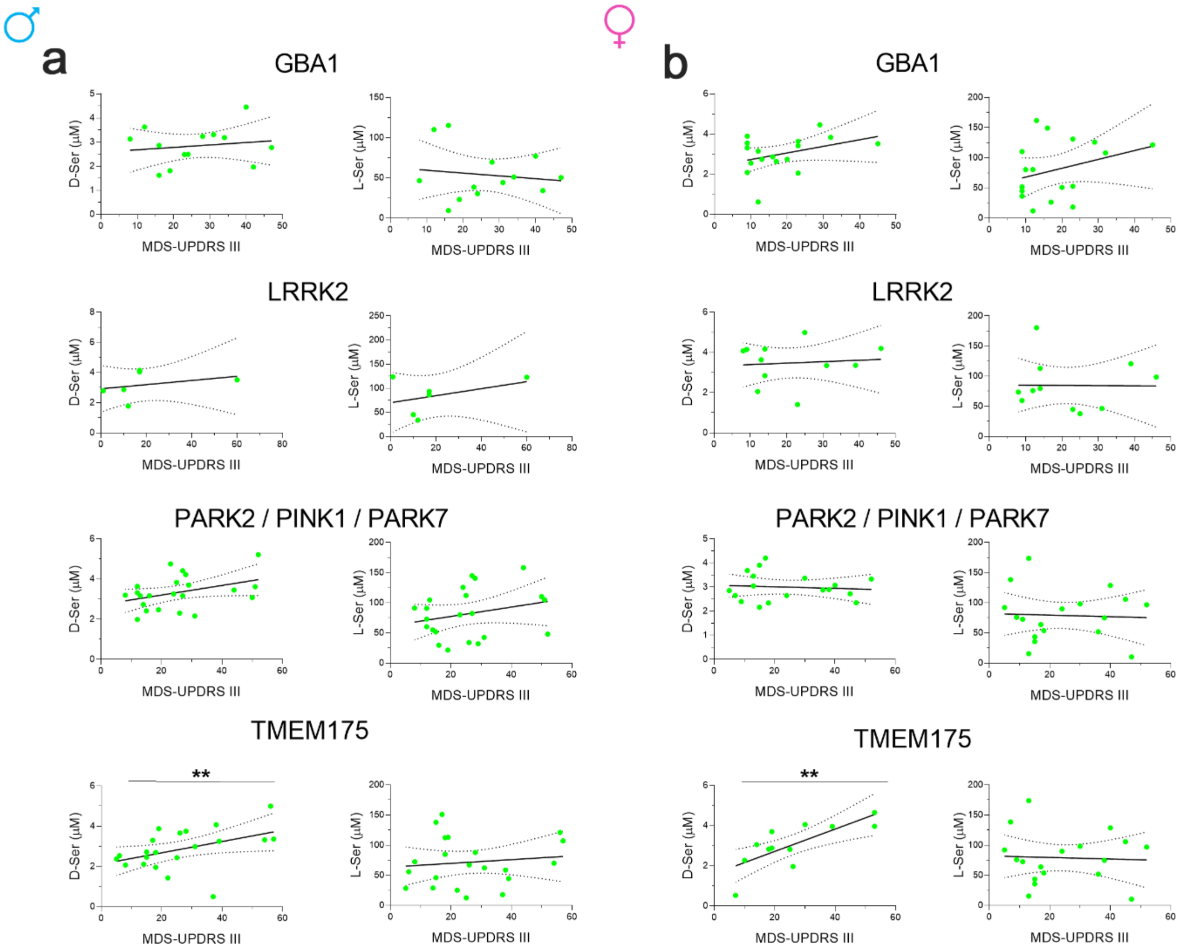


**Figure S5. Correlation of serum D-serine and L-serine concentrations with MDS-UPDRS III of genetic PD patients stratified for sex and for different pathogenic variants.** Scatterplots showing the association of D-serine (D-Ser) or L-serine (L-Ser) content with MDS-UPDRS III of male (a) and female (b) genetic PD stratified for GBA1, LRRK2, PARK2/PINK1/PARK7 and TMEM175 pathogenic variants. Best fit lines and 95% confidence intervals are shown. ** $p < 0.01$, Spearman's correlation test confirmed by partial correlation analysis.

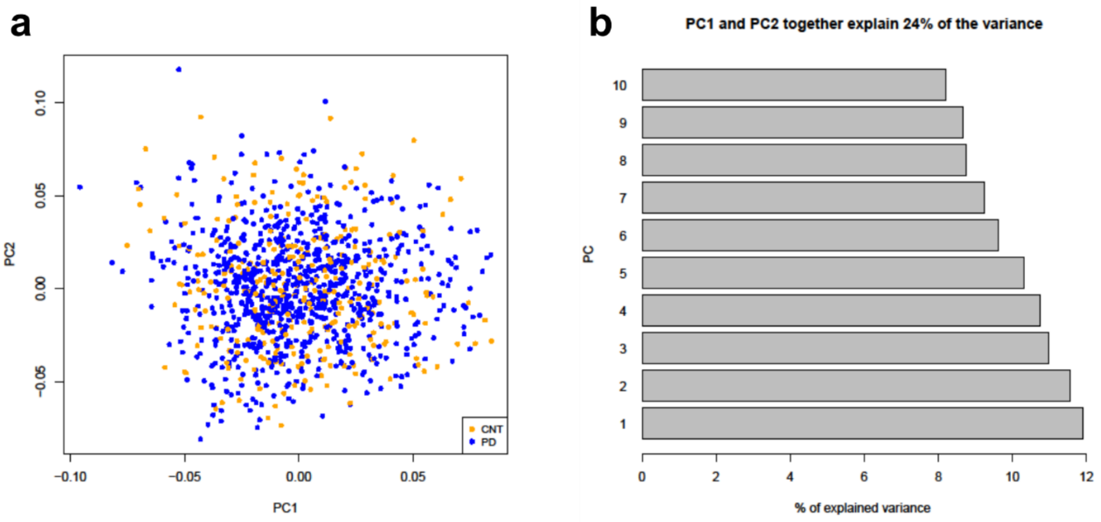


**Figure S6. Principal Component Analysis (PCA) of the Study Cohort. (a)** PCA analysis of the entire study Cohort (CNT_MNI, PD_MNI)**.** PCA Multidimensional Scaling plot was used to graphically show genetic diversity of the Cohort. **(b)** Graphical representation of PCs (1 to 10) contribution to the variance of the analyzed study cohort.

**Table S1. Pathogenic mutations in *LRRK2, GBA1, PARK2 (PRKN), PINK1, PARK7* and *TMEM175* in genetic-PD subtype**

| CHR | Genomic position (hg38) | dbSNP | Gene | RefSEq | Nucleotide Change | AA Change | Exonic Function | Cl | MAF gnomAD v 4.1 | CADD phred | PD patients N |
|---|---|---|---|---|---|---|---|---|---|---|---|
| 12 | 40340400 | rs34637584 | LRRK2 | NM_198578 | c.G6055A | p.G2019S | NSV | P | 0.0002721 | 35 | 14 |
| 12 | 40310434 | rs33939927 | LRRK2 | NM_198578 | c.C4321T | p.R1441C | NSV | P | 0.0000195 | 26.7 | 2 |
| 12 | 39859182 | rs34594498 | LRRK2 | NM_198578 | c.C1256T | p.A419V | NSV | P | 0.0001493 | 24.9 | 1 |
| 1 | 155266052 | rs76763715 | GBA1 | NM_000157 | c.A1226G | p.N409S | NSV | P | 0.001728 | 24.1 | 9 |
| 1 | 155266585 | rs2230288 | GBA1 | NM_000157 | c.G1093A | p.E365K | NSV | P | 0.01338 | 16.1 | 7 |
| 1 | 155296664 | rs75548401 | GBA1 | NM_000157 | c.C1223T | p.T408M | NSV | P | 0.008175 | 21.3 | 3 |
| 1 | 155265461 | rs421016 | GBA1 | NM_000157 | c.T1448C | p.L483P | NSV | P | 0.00007547 | 24.7 | 2 |
| 1 | 155267667 | rs367968666 | GBA1 | NM_000157 | c.T882G | p.H294Q | NSV | P | 0.0001678 | 12.25 | 2 |
| 1 | 155270905 | rs139626710 | GBA1 | NM_000157 | c.A49G | p.R17G | NSV | P | 0.00002119 | 8.03 | 1 |
| 1 | 155270204 | NA | GBA1 | NM_000157 | c.C198A | p.D66E | NSV | P | NA | 6.32 | 1 |
| 1 | 155268415 | rs381427 | GBA1 | NM_000157 | c.T689A | p.V230E | NSV | P | 0.00000339 | 22.3 | 1 |
| 1 | 155268386 | NA | GBA1 | NM_000157 | c.C718T | p.P240S | NSV | P | NA | 22.4 | 1 |
| 1 | 155266579 | NA | GBA1 | NM_000157 | c.C1099T | p.H367Y | NSV | P | NA | 25.7 | 1 |
| 1 | 155266470 | rs121908307 | GBA1 | NM_000157 | c.G1208C | p.S403T | NSV | P | 8.476e-7 | 22.8 | 1 |
| 1 | 155265999 | rs149171124 | GBA1 | NM_000157 | c.G1279A | p.E427K | NSV | P | 0.0002229 | 19.29 | 1 |
| 1 | 155265936 | rs1064651 | GBA1 | NM_000157 | c.G1342C | p.D448H | NSV | P | 0.0001043 | 23.3 | 1 |
| 1 | 155265414 | rs369068553 | GBA1 | NM_000157 | c.G1495C | p.V499L | NSV | P | 0.00004747 | 17.25 | 1 |
| 1 | 155265300 | NA | GBA1 | NM_000157 | c.G1515T | p.K505N | NSV | P | NA | 19.17 | 1 |
| 6 | 160929107 | rs775091228 | PRKN | NM_004562 | c.G1358A | p.W453X | Stop_gain | P | 0.00001186 | 44 | 2 |
| 6 | 161364788 | rs34424986 | PRKN | NM_004562 | c.C823T | p.R275W | NSV | P | 0.003732 | 26.1 | 2 |
| 6 | 160939137 | rs55830907 | PRKN | NM_004562 | c.C1204T | p.R402C | NSV | P | 0.001937 | 25.1 | 4 |
| 6 | 161841660 | rs55774500 | PRKN | NM_004562 | c.C245A | p.A82E | NSV | P | 0.002815 | 3.14 | 5 |
| 6 | 161633103 | rs9456735 | PRKN | NM_004562 | c.A574C | p.M192L | NSV | P | 0.0001356 | 20.7 | 1 |
| 6 | 161552303 | rs144032774 | PRKN | NM_004562 | c.G701A | p.R234Q | NSV | P | 0.0001280 | 20.9 | 1 |
| 6 | 161552285 | rs137853054 | PRKN | NM_004562 | c.C719T | p.T240M | NSV | P | 0.0001802 | 23.4 | 1 |
| 6 | 161552274 | NA | PRKN | NM_004562 | c.G730C | p.V244L | NSV | P | NA | 6.93 | 1 |
| 6 | 160939116 | NA | PRKN | NM_004562 | c.G1225T | p.E409X | Stop_gain | P | NA | 40 | 1 |
| 6 | 160939097 | rs778125254 | PRKN | NM_004562 | c.C1244A | p.T415N | NSV | P | 0.00000339 | 25.2 | 1 |
| 1 | 20311548 | rs138302371 | PINK1 | NM_032409 | c.C587T | p.P196L | NSV | P | 0.0003610 | 17.46 | 2 |
| 1 | 20322561 | rs74315356 | PINK1 | NM_032409 | c.G1311A | p.W437X | Stop_gain | P | 0.00000677 | 47 | 2 |
| 1 | 20311463 | rs768091663 | PINK1 | NM_032409 | c.G502C | p.A168P | NSV | P | 0.00000847 | 22.2 | 1 |
| 1 | 20311519 | rs143204084 | PINK1 | NM_032409 | c.G558C | p.K186N | NSV | P | 0.0002568 | 14.84 | 1 |
| 1 | 20318092 | NA | PINK1 | NM_032409 | c.C872A | p.A291D | NSV | P | NA | 26.3 | 1 |
| 1 | 20319083 | rs376323248 | PINK1 | NM_032409 | c.C976T | p.R326C | NSV | P | 0.00000508 | 26.9 | 1 |
| 1 | 20324025 | rs531477772 | PINK1 | NM_032409 | c.G1573A | p.D525N | NSV | P | 0.00009152 | 23.3 | 1 |
| 1 | 7910874 | rs71653619 | PARK7 | NM_007262 | c.G293A | p.R98Q | NSV | RF | 0.01043 | 20.9 | 14 |
| 1 | 7909346 | rs781094807 | PARK7 | NM_007262 | c.252+2->A | | splicing | N | 0.0002733 | NA | 2 |
| 4 | 958630 | rs752406714 | TMEM175 | NM_032326 | c.430_431del | p.V147Dfs*104 | Fs_del | P | 0.00000256 | NA | 4 |
| 4 | 964474 | rs565504915 | TMEM175 | NM_032326 | c.1281_1282del | p.A429Qfs*120 | Fs_del | P | 0.0003095 | NA | 3 |
| 4 | 958646 | NA | TMEM175 | NM_032326 | c.446_462del | p.A149Gfs*97 | Fs_del | P | NA | NA | 3 |
| 4 | 954054 | rs542936413 | TMEM175 | NM_032326 | c.C103T | p.R35C | NSV | P | 0.00002542 | 28.7 | 3 |
| 4 | 964406 | rs75307864 | TMEM175 | NM_032326 | c.C1213G | p.L405V | NSV | P | 0.003832 | 23.3 | 4 |
| 4 | 964433 | rs140597786 | TMEM175 | NM_032326 | c.C1240T | p.R414W | NSV | P | 0.004833 | 23.2 | 3 |
| 4 | 964197 | rs147762522 | TMEM175 | NM_032326 | c.G1004A | p.R335H | NSV | P | 0.001076 | 24.6 | 3 |
| 4 | 956673 | rs142778595 | TMEM175 | NM_032326 | c.T233C | p.I78T | NSV | P | 0.00005424 | 23.4 | 2 |

| | | | | | | | | | | | |
|---|---|---|---|---|---|---|---|---|---|---|---|
| 4 | 957441 | rs200834686 | TMEM175 | NM_032326 | c.A313G | p.T105A | NSV | P | 0.0001136 | 22.21 | 2 |
| 4 | 962038 | rs746980739 | TMEM175 | NM_032326 | c.C778T | p.R260C | NSV | P | 0.00004068 | 25.1 | 1 |
| 4 | 962068 | rs750645874 | TMEM175 | NM_032326 | c.G808A | p.A270T | NSV | P | 0.00000762 | 25.0 | 1 |
| 4 | 962100 | NA | TMEM175 | NM_032326 | c.C840G | p.I280M | NSV | P | NA | 24.7 | 1 |
| 4 | 964050 | rs778399444 | TMEM175 | NM_032326 | c.C857T | p.P286L | NSV | P | 0.00000254 | 23.9 | 1 |
| 4 | 964170 | rs148627215 | TMEM175 | NM_032326 | c.C977T | p.A326V | NSV | P | 0.00000169 | 7.41 | 1 |
| 4 | 964236 | rs147975675 | TMEM175 | NM_032326 | c.C1043T | p.S348L | NSV | P | 0.00005171 | 24.5 | 1 |
| 4 | 964464 | rs142744759 | TMEM175 | NM_032326 | c.C1271T | p.A424V | NSV | P | 0.00001188 | 22.6 | 1 |
| 4 | 964634 | rs201314478 | TMEM175 | NM_032326 | c.C1441T | p.R481W | NSV | P | 0.002678 | 10.6 | 1 |

CHR, Chromosome; hg38, human genome assembly 38; dbSNP, reference number in Single Nucleotide Polymorphism database; ref seq, reference number of the gene transcript; AA Change, amino acid change; CI, clinical interpretation; P, Pathogenic; RF, risk factor; N, novel; PD, Parkinson's disease; CADD phred, Combined Annotation Dependent Depletion; Fs_del, frameshift deletion; NSV, non-synonymous variant; MAF, Minor Allele Frequency, was referred to gnomAD v4.1 database; NA, Not Annotated.

**Table S2. Demographic and clinical features of male and female idiopathic and genetic PD patients.**

| Sex | Demographic and clinical characteristics | Idiopathic PD | | | | Genetic PD | | | | *p* value[a] |
|---|---|---|---|---|---|---|---|---|---|---|
| | | N | Median | IQR | | N | Median | IQR | | |
| Male | Age (years) | 65 | 67.0 | 61.0 | 74.0 | 64 | 68.0 | 61.5 | 72.0 | 0.938 |
| | Age at onset (years) | 65 | 61.0 | 55.0 | 66.0 | 64 | 60.0 | 55.5 | 65.5 | 0.869 |
| | Duration of disease (years) | 65 | 5.0 | 3.0 | 8.0 | 64 | 6.0 | 3.0 | 9.5 | 0.610 |
| | LEDD at interview (mg/die) | 65 | 400.0 | 300.0 | 560.0 | 64 | 451.0 | 320.0 | 724.0 | 0.304 |
| | MDS-UPDRS III | 65 | 24.0 | 13.0 | 35.0 | 64 | 22.5 | 14.5 | 31.0 | 0.668 |
| Female | Age (years) | 56 | 69.5 | 62.5 | 74.0 | 60 | 68.0 | 63.5 | 73.5 | 0.560 |
| | Age at onset (years) | 56 | 64.0 | 56.5 | 69.0 | 60 | 61.5 | 52.5 | 65.0 | 0.042 |
| | Duration of disease (years) | 56 | 5.0 | 3.0 | 7.0 | 60 | 7.0 | 3.5 | 12.0 | 0.012 |
| | LEDD at interview (mg/die) | 56 | 400.0 | 302.5 | 600.0 | 60 | 483.0 | 300.0 | 645.0 | 0.730 |
| | MDS-UPDRS III | 56 | 21.0 | 13.0 | 28.5 | 60 | 17.5 | 12.0 | 30.0 | 0.525 |

Abbreviations: N, number of subjects; IQR, interquartile range; LEDD, Levodopa equivalent daily dose; MDS-UPDRS III, Movement Disorders Society Unified Parkinson's Disease Rating Scale, part III.
[a] Mann-Whitney U test.

**Table S3. Serum levels of D- and L- amino acids in Parkinson's disease and healthy controls groups.**

| Amino acids | Healthy controls | | | | Parkinson's disease | | | | Mann-Whitney | ANCOVA[a] | | % of HC |
|---|---|---|---|---|---|---|---|---|---|---|---|---|
| | N | Median | IQR | | N | Median | IQR | | *p value* | $F_{(1;443)}$ | *p* value | |
| L-aspartate (µM) | 203 | 11.0 | 7.9 | 16.0 | 245 | 10.2 | 7.5 | 13.3 | **0.014** | 3.909 | **0.049** | -8 |
| L-glutamate (µM) | 203 | 26.2 | 18.0 | 39.2 | 245 | 22.7 | 14.9 | 32.8 | **0.005** | 5.628 | **0.018** | -13 |
| L-asparagine (µM) | 203 | 20.1 | 12.0 | 32.2 | 245 | 17.9 | 10.7 | 26.7 | **0.032** | 2.862 | 0.091 | |
| D-serine (µM) | 203 | 2.7 | 2.0 | 3.1 | 245 | 2.8 | 2.3 | 3.4 | **0.011** | 0.138 | 0.711 | |
| L-serine (µM) | 203 | 74.4 | 43.0 | 106.4 | 245 | 64.6 | 41.0 | 96.7 | 0.069 | | | |
| D-serine/total serine (%) | 203 | 3.3 | 2.3 | 5.1 | 245 | 4.0 | 3.0 | 5.9 | **<0.0001** | 4.383 | **0.037** | 21 |
| L-glutamine (µM) | 203 | 173.7 | 108.0 | 234.0 | 245 | 160.5 | 105.8 | 230.4 | 0.371 | | | |
| Glycine (µM) | 203 | 144.0 | 104.2 | 196.1 | 245 | 141.3 | 98.9 | 205.4 | 0.986 | | | |
| L-glutamine/ L-glutamate | 203 | 6.2 | 4.6 | 7.7 | 245 | 6.8 | 5.3 | 8.4 | **0.001** | 5.033 | **0.025** | 10 |

Abbreviations: N, number of subjects; IQR, interquartile range. Significant p values are shown in bold.

[a]Age-, sex- and LEDD-adjusted.

**Table S4. Serum levels of D- and L-amino acids in male and female Parkinson's disease patients and healthy controls.**

| | Healthy controls | | | | Parkinson's disease | | | | Mann-Whitney | ANCOVA[a] | | % of HC |
|---|---|---|---|---|---|---|---|---|---|---|---|---|
| **Male** | N | Median | IQR | | N | Median | IQR | | p value | $F_{(1;206)}$ | p value | |
| L-aspartate (µM) | 80 | 12.2 | 8.8 | 16.7 | 129 | 9.7 | 6.8 | 11.8 | **<0.0001** | 13.582 | **0.0003** | -21 |
| L-glutamate (µM) | 80 | 31 | 21.8 | 44 | 129 | 22.0 | 14.2 | 31.4 | **<0.0001** | 11.553 | **0.0008** | -29 |
| L-asparagine (µM) | 80 | 22.7 | 15.2 | 34.5 | 129 | 16.1 | 9.6 | 24.9 | **0.0002** | 11.400 | **0.0009** | -29 |
| D-serine (µM) | 80 | 2.8 | 2.4 | 3.2 | 129 | 2.7 | 2.2 | 3.2 | 0.482 | 1.536 | 0.217 | |
| L-serine (µM) | 80 | 75.2 | 53 | 107 | 129 | 56.8 | 33.9 | 84.7 | **0.0004** | 9.877 | **0.002** | -24 |
| D-serine/total serine (%) | 80 | 3.5 | 2.4 | 4.8 | 129 | 4.6 | 3.2 | 6.3 | **0.0003** | 7.976 | **0.005** | 30 |
| L-glutamine (µM) | 80 | 183.5 | 136.1 | 240.2 | 129 | 144.1 | 97.3 | 202.1 | **0.003** | 8.046 | **0.005** | -21 |
| Glycine (µM) | 80 | 151.3 | 108.5 | 201.8 | 129 | 124.8 | 90.7 | 179.5 | **0.014** | 7.632 | **0.006** | -18 |
| L-glutamine/ L-glutamate | 80 | 6.1 | 4.4 | 7.3 | 129 | 6.5 | 5.0 | 8.1 | **0.031** | 0.726 | 0.395 | |
| **Female** | N | Median | IQR | | N | Median | IQR | | p value | $F_{(1;237)}$ | p value | % of HC |
| L-aspartate (µM) | 123 | 10.5 | 7.3 | 15.2 | 116 | 11.2 | 8.5 | 15.4 | 0.422 | | | |
| L-glutamate (µM) | 123 | 23.5 | 14.8 | 35.4 | 116 | 23.3 | 15.4 | 33.2 | 0.989 | | | |
| L-asparagine (µM) | 123 | 18.4 | 9.9 | 30.2 | 116 | 20.4 | 12.0 | 27.9 | 0.609 | | | |
| D-serine (µM) | 123 | 2.4 | 1.9 | 3 | 116 | 2.9 | 2.3 | 3.5 | **0.0002** | 1.551 | 0.214 | |
| L-serine (µM) | 123 | 66.9 | 35.6 | 105.4 | 116 | 76.0 | 46.9 | 107.9 | 0.404 | | | |
| D-serine/total serine (%) | 123 | 3.2 | 2.3 | 5.4 | 116 | 3.6 | 2.9 | 5.3 | 0.061 | | | |
| L-glutamine (µM) | 123 | 155.3 | 94.1 | 218.4 | 116 | 185.9 | 122.4 | 241.2 | 0.122 | | | |
| Glycine (µM) | 123 | 143.6 | 95.6 | 195.9 | 116 | 171.1 | 115.1 | 227.5 | **0.009** | 1.654 | 0.200 | |
| L-glutamine/ L-glutamate | 123 | 6.2 | 4.7 | 7.9 | 116 | 7.2 | 5.8 | 8.8 | **0.005** | 5.283 | **0.022** | 17 |

Abbreviations: N, number of subjects; IQR, interquartile range. Significant p values are shown in bold.

[a]Age-, and LEDD-adjusted.

### Table S5. Serum levels of D- and L-amino acids in idiopathic PD, genetic PD and healthy controls.

| Amino acids | Healthy controls (N=203) | | | Idiopathic PD (N=121) | | | | Genetic PD (N=124) | | | | Kruskal-Wallis | ANCOVA[a] | | Dunn's test with Bonferroni correction | | |
|---|---|---|---|---|---|---|---|---|---|---|---|---|---|---|---|---|---|
| | Median | IQR | | Median | IQR | | % of HC | Median | IQR | | % of HC | *p value* | $F_{(2;443)}$ | *p value* | *HC vs Idiopathic PD* | *HC vs genetic PD* | *Idiopathic PD vs genentic PD* |
| L-aspartate (µM) | 11.0 | 7.9 | 16.0 | 9.9 | 6.9 | 12.8 | -10 | 10.4 | 8.0 | 13.9 | -6 | **0.012** | 3.450 | **0.033** | **0.009** | 0.779 | 0.29 |
| L-glutamate (µM) | 26.2 | 18.0 | 39.2 | 21.0 | 13.4 | 29.6 | -20 | 25.0 | 16.3 | 33.6 | -5 | **0.001** | 6.613 | **0.001** | **<0.0001** | >0.999 | **0.027** |
| L-asparagine (µM) | 20.1 | 12.0 | 32.2 | 17.1 | 10.4 | 24.6 | -15 | 20.1 | 11.8 | 28.2 | 0 | **0.015** | 3.313 | **0.037** | **0.012** | >0.999 | 0.147 |
| D-serine (µM) | 2.7 | 2.0 | 3.1 | 2.6 | 2.1 | 3.0 | -3 | 3.1 | 2.5 | 3.6 | 16 | **<0.0001** | 5.089 | **0.007** | >0.999 | **<0.0001** | **<0.0001** |
| L-serine (µM) | 74.4 | 43.0 | 106.4 | 58.6 | 35.5 | 85.0 | -21 | 72.7 | 44.9 | 110.1 | -2 | **0.010** | 3.818 | **0.023** | **0.012** | >0.999 | **0.046** |
| D-serine/total serine (%) | 3.3 | 2.3 | 5.1 | 3.9 | 3.2 | 5.9 | 17 | 4.2 | 2.9 | 5.9 | 26 | **0.000** | 2.314 | 0.1 | | | |
| L-glutamine (µM) | 173.7 | 108.0 | 234.0 | 155.3 | 97.7 | 217.0 | -11 | 165.4 | 110.8 | 243.0 | -5 | 0.176 | | | | | |
| Glycine (µM) | 144.0 | 104.2 | 196.1 | 124.8 | 91.4 | 185.3 | -13 | 157.8 | 106.8 | 223.0 | 10 | **0.017** | 3.847 | **0.022** | 0.312 | 0.349 | **0.013** |
| L-glutamine/ L-glutamate | 6.2 | 4.6 | 7.7 | 7.0 | 5.5 | 9.1 | 14 | 6.7 | 5.3 | 8.0 | 7 | **0.002** | 4.283 | **0.014** | **0.002** | 0.166 | 0.479 |

Abbreviations: N. number of subjects; IQR. interquartile range. Significant p values are shown in bold.
[a]Age-. sex- and LEDD-adjusted.

**Table S6**. **Serum levels of D- and L-amino acids in male idiopathic PD, genetic PD and healthy controls.**

| Male | HC (N=80) | | | Idiopathic PD (N=65) | | | | Genetic PD (N=64) | | | | Kruskal-Wallis | ANCOVA[a] | | Dunn's test with Bonferroni correction | | |
|---|---|---|---|---|---|---|---|---|---|---|---|---|---|---|---|---|---|
| Amino acids (μM) | Median | IQR | | Median | IQR | | % of HC | Median | IQR | | % of HC | *p value* | $F_{(2;206)}$ | *p-value* | *HC vs Idiopathic PD* | *HC vs genetic PD* | *Idiopathic PD vs genetic PD* |
| L-aspartate (μM) | 12.2 | 8.8 | 16.7 | 8.9 | 6.3 | 10.8 | -27.10 | 9.9 | 7.4 | 12.7 | -19.1 | **<0.0001** | 8.788 | **0.0002** | **<0.0001** | **0.028** | 0.6833 |
| L-glutamate (μM) | 31.0 | 21.8 | 44.0 | 19.5 | 12.5 | 28.1 | -37.31 | 26.3 | 17.0 | 34.3 | -15.2 | **<0.0001** | 10.150 | **<0.0001** | **<0.0001** | 0.062 | **0.021** |
| L-asparagine (μM) | 22.7 | 15.2 | 34.5 | 14.8 | 8.5 | 20.8 | -34.90 | 18.7 | 10.8 | 27.2 | -17.5 | **<0.0001** | 8.409 | **0.0003** | **<0.0001** | 0.132 | 0.073 |
| D-serine (μM) | 2.8 | 2.4 | 3.2 | 2.5 | 2.0 | 2.9 | -13.16 | 3.1 | 2.5 | 3.6 | 11.3 | **0.0003** | 7.455 | **0.0010** | **0.017** | 0.328 | **<0.0001** |
| L-serine (μM) | 75.2 | 53.0 | 107.0 | 52.6 | 26.8 | 71.7 | -29.97 | 68.4 | 43.4 | 106.0 | -9.1 | **<0.0001** | 9.421 | **0.0001** | **<0.0001** | 0.42 | **0.013** |
| D-serine/total serine (%) | 3.5 | 2.4 | 4.8 | 4.2 | 3.4 | 7.5 | 20.30 | 4.6 | 3.0 | 6.0 | 32.2 | **0.001** | 4.638 | **0.0110** | **0.002** | **0.021** | >0.9999 |
| L-glutamine (μM) | 183.5 | 136.1 | 240.2 | 132.8 | 90.0 | 181.5 | -27.63 | 166.7 | 105.0 | 238.7 | -9.2 | **0.001** | 6.401 | **0.0020** | **0.001** | 0.508 | 0.079 |
| Glycine (μM) | 151.3 | 108.5 | 201.8 | 111.6 | 78.5 | 149.1 | -26.19 | 142.7 | 97.7 | 211.0 | -5.7 | **0.0002** | 8.877 | **0.0002** | **<0.0001** | >0.999 | **0.003** |
| L-glutamine/ L-glutamate | 6.1 | 4.4 | 7.3 | 6.5 | 5.1 | 9.0 | 7.43 | 6.3 | 5.0 | 7.8 | 4.2 | 0.072 | | | | | |

Abbreviations: N. number of subjects; IQR. interquartile range. Significant p values are shown in bold.
[a]Age- and LEDD-adjusted

**Table S7. Serum levels of D- and L-amino acids in female idiopathic PD, genetic PD and healthy controls.**

| **Female** | HC (N=123) | | | Idiopathic PD (N=56) | | | | Genetic PD (N=60) | | | | Kruscal -Wallis | ANCOVA[a] | | Dunn's test with Bonferroni correction | | |
|---|---|---|---|---|---|---|---|---|---|---|---|---|---|---|---|---|---|
| Amino acids (µM) | Median | IQR | | Median | IQR | | % of HC | Median | IQR | | % of HC | *p value* | $F_{(2;237)}$ | *p-value* | *HC vs Idiopathic PD* | *HC vs genetic PD* | *Idiopathic PD vs genetic PD* |
| L-aspartate (µM) | 10.5 | 7.3 | 15.2 | 11.2 | 8.9 | 14.7 | 6.24 | 11.2 | 8.2 | 15.7 | 6.7 | 0.673 | 0.275 | 0.759 | | | |
| L-glutamate (µM) | 23.5 | 14.8 | 35.4 | 23.0 | 14.8 | 32.5 | -2.14 | 23.6 | 15.8 | 33.4 | 0.2 | 0.68 | 0.594 | 0.553 | | | |
| L-asparagine (µM) | 18.4 | 9.9 | 30.2 | 20.0 | 12.1 | 27.3 | 8.63 | 22.2 | 12.0 | 30.7 | 20.5 | 0.766 | 0.393 | 0.676 | | | |
| D-serine (µM) | 2.4 | 1.9 | 3.0 | 2.7 | 2.2 | 3.4 | 9.74 | 3.0 | 2.6 | 3.7 | 25.7 | **<0.0001** | 3.0 | 0.065 | | | |
| L-serine (µM) | 66.9 | 35.6 | 105.4 | 77.6 | 45.7 | 97.8 | 15.90 | 75.9 | 46.9 | 115.5 | 13.3 | 0.611 | 0.255 | 0.775 | | | |
| D-serine/total serine (%) | 3.2 | 2.3 | 5.4 | 3.5 | 3.0 | 4.4 | 8.16 | 3.7 | 2.8 | 5.8 | 15.8 | 0.165 | 0.123 | 0.884 | | | |
| L-glutamine (µM) | 155.3 | 94.1 | 218.4 | 188.5 | 129.3 | 231.3 | 21.38 | 164.4 | 119.6 | 247.4 | 5.9 | 0.303 | 0.758 | 0.47 | | | |
| Glycine (µM) | 143.6 | 95.6 | 195.9 | 173.8 | 110.6 | 216.7 | 21.06 | 169.6 | 122.8 | 233.2 | 18.1 | **0.026** | 0.928 | 0.397 | | | |
| L-glutamine/ L-glutamate | 6.2 | 4.7 | 7.9 | 7.4 | 5.8 | 9.6 | 18.25 | 7.0 | 5.6 | 8.1 | 12.1 | **0.009** | 3,971 | **0.02** | **0.008** | 0.328 | 0.638 |

Abbreviations: N. number of subjects; IQR. interquartile range. Significant p values are shown in bold.

[a]Age- and LEDD-adjusted

**Table S8. Serum levels of D- and L-amino acids in male and female idiopathic PD and genetic PD.**

| | Idiopathic PD | | | Genetic PD | | | Mann-Whitney | ANCOVA[a] | |
|---|---|---|---|---|---|---|---|---|---|
| **Male** | (N=65) | | | (N=64) | | | | | |
| Amino acids | Median | IQR | | Median | IQR | | *p value* | $F_{(1;124)}$ | *p value* |
| L-aspartate (µM) | 8.9 | 6.3 | 10.8 | 9.9 | 7.4 | 12.7 | **0.032** | 3.985 | **0.048** |
| L-glutamate (µM) | 19.5 | 12.5 | 28.1 | 26.3 | 17.0 | 34.3 | **0.006** | 7.582 | **0.007** |
| L-asparagine (µM) | 14.8 | 8.5 | 20.8 | 18.7 | 10.8 | 27.2 | **0.021** | 5.255 | **0.024** |
| D-serine (µM) | 2.5 | 2.0 | 2.9 | 3.1 | 2.5 | 3.6 | **0.000** | 10.587 | **0.001** |
| L-serine (µM) | 52.6 | 26.8 | 71.7 | 68.4 | 43.4 | 106.0 | **0.005** | 8.051 | **0.005** |
| D-serine/total serine (%) | 4.2 | 3.4 | 7.5 | 4.6 | 3.0 | 6.0 | 0.457 | 1.440 | 0.232 |
| L-glutamine (µM) | 132.8 | 90.0 | 181.5 | 166.7 | 105.0 | 238.7 | **0.027** | 4.442 | **0.037** |
| Glycine (µM) | 111.6 | 78.5 | 149.1 | 142.7 | 97.7 | 211.0 | **0.001** | 9.462 | **0.003** |
| L-glutamine/ L-glutamate | 6.5 | 5.1 | 9.0 | 6.3 | 5.0 | 7.8 | 0.412 | 1.056 | 0.306 |

| **Female** | (N=56) | | | (N=60) | | | | | |
|---|---|---|---|---|---|---|---|---|---|
| Amino acids | Median | IQR | | Median | IQR | | *p value* | $F_{(1;111)}$ | *p value* |
| L-aspartate (µM) | 11.2 | 8.9 | 14.7 | 11.2 | 8.2 | 15.7 | 0.715 | | |
| L-glutamate (µM) | 23.0 | 14.8 | 32.5 | 23.6 | 15.8 | 33.4 | 0.410 | | |
| L-asparagine (µM) | 20.0 | 12.1 | 27.3 | 22.2 | 12.0 | 30.7 | 0.607 | | |
| D-serine (µM) | 2.7 | 2.2 | 3.4 | 3.0 | 2.6 | 3.7 | 0.033 | 1.436 | 0.233 |
| L-serine (µM) | 77.6 | 45.7 | 97.8 | 75.9 | 46.9 | 115.5 | 0.558 | | |
| D-serine/total serine (%) | 3.5 | 3.0 | 4.4 | 3.7 | 2.8 | 5.8 | 0.736 | | |
| L-glutamine (µM) | 188.5 | 129.3 | 231.3 | 164.4 | 119.6 | 247.4 | 0.916 | | |
| Glycine (µM) | 173.8 | 110.6 | 216.7 | 169.6 | 122.8 | 233.2 | 0.536 | | |
| L-glutamine/ L-glutamate | 7.4 | 5.8 | 9.6 | 7.0 | 5.6 | 8.1 | 0.189 | | |

Abbreviations: N.number of subjects; IQR. interquartile range. Significant p values are shown in bold.

[a]Age-, LEDD- and disease duration-adjusted

**Table S9**. **Linear model results: group × sex interaction on serum NMDAR-related amino acid levels (full cohort). Results from ordinary least squares (OLS) linear models fitted separately for each of the nine amino acid outcomes.**

| Outcome | Term | β | SE | t | p (raw) | p (FDR) |
|---|---|---|---|---|---|---|
| **L-Asp** | Intercept | 2.367 | 0.143 | 16.612 | < 0.001 | |
| | Group (PD vs HC) | 0.041 | 0.062 | 0.655 | 0.5125 | |
| | Sex (M vs F) | 0.076 | 0.065 | 1.175 | 0.2406 | |
| | Age | 0.000 | 0.002 | 0.072 | 0.9427 | |
| | **Group × Sex (interaction)** | -0.287 | 0.084 | -3.428 | < 0.001 | **0.0026** |
| **L-Glu** | Intercept | 2.973 | 0.186 | 16.021 | < 0.001 | |
| | Group (PD vs HC) | -0.007 | 0.081 | -0.087 | 0.9310 | |
| | Sex (M vs F) | 0.237 | 0.079 | 2.984 | 0.0030 | |
| | Age | 0.003 | 0.003 | 0.937 | 0.3495 | |
| | **Group × Sex (interaction)** | -0.333 | 0.108 | -3.089 | 0.0021 | **0.0031** |

| Outcome | Term | β | SE | t | p (raw) | p (FDR) |
|---|---|---|---|---|---|---|
| **L-Asn** | Intercept | 2.834 | 0.214 | 13.243 | < 0.001 | |
| | Group (PD vs HC) | 0.061 | 0.091 | 0.667 | 0.5048 | |
| | Sex (M vs F) | 0.210 | 0.097 | 2.160 | 0.0313 | |
| | Age | -0.000 | 0.003 | -0.030 | 0.9760 | |
| | **Group × Sex (interaction)** | -0.388 | 0.127 | -3.052 | 0.0024 | **0.0031** |
| **D-Ser** | Intercept | 0.493 | 0.119 | 4.138 | < 0.001 | |
| | Group (PD vs HC) | 0.151 | 0.051 | 2.963 | 0.0032 | |
| | Sex (M vs F) | 0.156 | 0.050 | 3.138 | 0.0018 | |
| | Age | 0.006 | 0.002 | 3.136 | 0.0018 | |
| | **Group × Sex (interaction)** | -0.217 | 0.068 | -3.203 | 0.0015 | **0.0026** |
| **L-Ser** | Intercept | 4.174 | 0.213 | 19.591 | < 0.001 | |
| | Group (PD vs HC) | 0.097 | 0.090 | 1.082 | 0.2797 | |

| Outcome | Term | β | SE | t | p (raw) | p (FDR) |
|---|---|---|---|---|---|---|
| | Sex (M vs F) | 0.161 | 0.092 | 1.750 | 0.0809 | |
| | Age | -0.001 | 0.003 | -0.351 | 0.7259 | |
| | **Group × Sex (interaction)** | -0.406 | 0.125 | -3.260 | 0.0012 | **0.0026** |
| **D-Ser/tot-Ser (ratio)** | Intercept | 0.895 | 0.183 | 4.893 | < 0.001 | |
| | Group (PD vs HC) | 0.054 | 0.076 | 0.710 | 0.4779 | |
| | Sex (M vs F) | -0.003 | 0.082 | -0.033 | 0.9736 | |
| | Age | 0.006 | 0.003 | 2.213 | 0.0274 | |
| | **Group × Sex (interaction)** | 0.177 | 0.106 | 1.665 | 0.0966 | 0.1087 |
| **L-Gln** | Intercept | 4.939 | 0.178 | 27.763 | < 0.001 | |
| | Group (PD vs HC) | 0.119 | 0.074 | 1.609 | 0.1083 | |
| | Sex (M vs F) | 0.177 | 0.080 | 2.218 | 0.0271 | |
| | Age | 0.001 | 0.003 | 0.219 | 0.8270 | |

| Outcome | Term | β | SE | t | p (raw) | p (FDR) |
|---|---|---|---|---|---|---|
| | **Group × Sex (interaction)** | -0.338 | 0.104 | -3.243 | 0.0013 | **0.0026** |
| **Gly** | Intercept | 4.845 | 0.164 | 29.542 | < 0.001 | |
| | Group (PD vs HC) | 0.181 | 0.066 | 2.733 | 0.0065 | |
| | Sex (M vs F) | 0.062 | 0.070 | 0.876 | 0.3816 | |
| | Age | 0.001 | 0.003 | 0.376 | 0.7072 | |
| | **Group × Sex (interaction)** | -0.358 | 0.095 | -3.777 | < 0.001 | **0.0016** |
| **L-Gln/L-Glu (ratio)** | Intercept | 1.967 | 0.137 | 14.324 | < 0.001 | |
| | Group (PD vs HC) | 0.126 | 0.048 | 2.603 | 0.0095 | |
| | Sex (M vs F) | -0.059 | 0.050 | -1.187 | 0.2359 | |
| | Age | -0.002 | 0.002 | -1.011 | 0.3127 | |
| | **Group × Sex (interaction)** | -0.005 | 0.069 | -0.074 | 0.9413 | 0.9413 |

Model formula: log(amino acid) ~ group + sex + age + group:sex, where group = HC (0) vs PD (1) and sex = female (0) vs male (1). Coefficients (β), robust standard errors (SE), t-statistics and raw p-values are reported for all model terms. The p (FDR) column reports Benjamini-Hochberg false discovery rate-corrected p-values for the interaction term only, derived from correction across the 9 interaction tests. Bold and highlighted cells in the p (FDR) column indicate significance at $p < 0.05$ after FDR correction. All inference is based on heteroscedasticity-consistent (HC3) robust standard errors. n = 443 (5 subjects excluded due to missing age data).

**Model:** log(amino acid) ~ group + sex + age + group:sex. group: 0 = HC, 1 = PD; sex: 0 = female, 1 = male. Inference based on heteroscedasticity-consistent robust standard errors (HC3). p (FDR): Benjamini-Hochberg correction applied to the 9 interaction p-values. Bold/shaded cells in p (FDR) column indicate significance at $p < 0.05$ after FDR correction. n = 443 (5 subjects excluded due to missing age).

**Table S10**. **Linear model results: subtype × sex interaction on serum NMDAR-related amino acid levels (PD patients only).**

| Outcome | Term | β | SE | t | p (raw) | p (FDR) |
|---|---|---|---|---|---|---|
| **L-Asp** | Intercept | 2.382 | 0.232 | 10.247 | < 0.001 | |
| | Subtype (gPD vs iPD) | 0.045 | 0.083 | 0.545 | 0.5861 | |
| | Sex (M vs F) | -0.257 | 0.073 | -3.524 | < 0.001 | |
| | Age | 0.000 | 0.003 | 0.030 | 0.9759 | |
| | Disease duration | -0.002 | 0.004 | -0.491 | 0.6242 | |
| | LEDD | 0.000 | 0.000 | 0.562 | 0.5744 | |
| | **Subtype × Sex (interaction)** | 0.097 | 0.106 | 0.913 | 0.3621 | 0.4073 |
| **L-Glu** | Intercept | 3.020 | 0.322 | 9.368 | < 0.001 | |
| | Subtype (gPD vs iPD) | 0.124 | 0.110 | 1.133 | 0.2582 | |
| | Sex (M vs F) | -0.169 | 0.100 | -1.691 | 0.0922 | |
| | Age | 0.001 | 0.004 | 0.244 | 0.8075 | |
| | Disease duration | -0.004 | 0.006 | -0.622 | 0.5346 | |

| Outcome | Term | β | SE | t | p (raw) | p (FDR) |
|---|---|---|---|---|---|---|
| | LEDD | 0.000 | 0.000 | 0.380 | 0.7043 | |
| | **Subtype × Sex (interaction)** | 0.147 | 0.145 | 1.012 | 0.3126 | 0.4019 |
| **L-Asn** | Intercept | 2.742 | 0.335 | 8.189 | < 0.001 | |
| | Subtype (gPD vs iPD) | 0.053 | 0.124 | 0.429 | 0.6682 | |
| | Sex (M vs F) | -0.278 | 0.116 | -2.399 | 0.0172 | |
| | Age | 0.002 | 0.005 | 0.334 | 0.7387 | |
| | Disease duration | -0.001 | 0.009 | -0.157 | 0.8755 | |
| | LEDD | 0.000 | 0.000 | 0.382 | 0.7025 | |
| | **Subtype × Sex (interaction)** | 0.209 | 0.165 | 1.267 | 0.2063 | 0.3713 |
| **D-Ser** | Intercept | 0.661 | 0.158 | 4.198 | < 0.001 | |
| | Subtype (gPD vs iPD) | 0.072 | 0.065 | 1.114 | 0.2662 | |
| | Sex (M vs F) | -0.124 | 0.058 | -2.135 | 0.0337 | |

| Outcome | Term | β | SE | t | p (raw) | p (FDR) |
|---|---|---|---|---|---|---|
| | Age | 0.004 | 0.002 | 1.926 | 0.0553 | |
| | Disease duration | 0.002 | 0.004 | 0.415 | 0.6784 | |
| | LEDD | 0.000 | 0.000 | 0.664 | 0.5076 | |
| | **Subtype × Sex (interaction)** | 0.135 | 0.090 | 1.493 | 0.1369 | 0.3226 |
| **L-Ser** | Intercept | 4.053 | 0.347 | 11.696 | < 0.001 | |
| | Subtype (gPD vs iPD) | 0.037 | 0.124 | 0.302 | 0.7627 | |
| | Sex (M vs F) | -0.386 | 0.115 | -3.366 | < 0.001 | |
| | Age | 0.001 | 0.005 | 0.294 | 0.7691 | |
| | Disease duration | -0.005 | 0.008 | -0.604 | 0.5466 | |
| | LEDD | 0.000 | 0.000 | 0.996 | 0.3204 | |
| | **Subtype × Sex (interaction)** | 0.295 | 0.167 | 1.769 | 0.0782 | 0.3226 |
| **D-Ser/tot-Ser (ratio)** | Intercept | 1.176 | 0.296 | 3.972 | < 0.001 | |

| Outcome | Term | β | SE | t | p (raw) | p (FDR) |
|---|---|---|---|---|---|---|
| | Subtype (gPD vs iPD) | 0.032 | 0.098 | 0.331 | 0.7409 | |
| | Sex (M vs F) | 0.245 | 0.097 | 2.542 | 0.0117 | |
| | Age | 0.003 | 0.004 | 0.653 | 0.5144 | |
| | Disease duration | 0.006 | 0.006 | 0.912 | 0.3626 | |
| | LEDD | -0.000 | 0.000 | -0.726 | 0.4684 | |
| | **Subtype × Sex (interaction)** | -0.147 | 0.134 | -1.092 | 0.2760 | 0.4019 |
| **L-Gln** | Intercept | 4.948 | 0.279 | 17.719 | < 0.001 | |
| | Subtype (gPD vs iPD) | 0.008 | 0.097 | 0.082 | 0.9347 | |
| | Sex (M vs F) | -0.255 | 0.095 | -2.695 | 0.0075 | |
| | Age | 0.002 | 0.004 | 0.457 | 0.6480 | |
| | Disease duration | -0.000 | 0.007 | -0.019 | 0.9848 | |
| | LEDD | 0.000 | 0.000 | 0.626 | 0.5317 | |
| | **Subtype × Sex (interaction)** | 0.198 | 0.135 | 1.468 | 0.1434 | 0.3226 |

| Outcome | Term | β | SE | t | p (raw) | p (FDR) |
|---|---|---|---|---|---|---|
| **Gly** | Intercept | 4.971 | 0.280 | 17.774 | < 0.001 | |
| | Subtype (gPD vs iPD) | 0.039 | 0.095 | 0.413 | 0.6797 | |
| | Sex (M vs F) | -0.403 | 0.090 | -4.501 | < 0.001 | |
| | Age | 0.000 | 0.004 | 0.119 | 0.9051 | |
| | Disease duration | -0.000 | 0.005 | -0.034 | 0.9726 | |
| | LEDD | 0.000 | 0.000 | 1.499 | 0.1353 | |
| | **Subtype × Sex (interaction)** | 0.231 | 0.127 | 1.818 | 0.0703 | 0.3226 |
| **L-Gln/L-Glu (ratio)** | Intercept | 1.927 | 0.208 | 9.265 | < 0.001 | |
| | Subtype (gPD vs iPD) | -0.116 | 0.069 | -1.695 | 0.0914 | |
| | Sex (M vs F) | -0.086 | 0.068 | -1.262 | 0.2083 | |
| | Age | 0.001 | 0.003 | 0.245 | 0.8070 | |
| | Disease duration | 0.004 | 0.004 | 0.869 | 0.3859 | |

| Outcome | Term | β | SE | t | p (raw) | p (FDR) |
|---|---|---|---|---|---|---|
| | LEDD | 0.000 | 0.000 | 0.077 | 0.9384 | |
| | **Subtype × Sex (interaction)** | 0.050 | 0.093 | 0.541 | 0.5893 | 0.5893 |

Results from ordinary least squares (OLS) linear models fitted separately for each of the nine amino acid outcomes in PD patients only. Model formula: log(amino acid) ~ subtype + sex + age + disease duration + LEDD + subtype:sex, where subtype = iPD (0) vs gPD (1) and sex = female (0) vs male (1). Coefficients (β), robust standard errors (SE), t-statistics and raw p-values are reported for all model terms. The p (FDR) column reports Benjamini-Hochberg false discovery rate-corrected p-values for the interaction term only, derived from correction across the 9 interaction tests. No interaction term reached significance after FDR correction. All inference is based on heteroscedasticity-consistent (HC3) robust standard errors. n = 245.

**Model:** log(amino acid) ~ subtype + sex + age + disease duration + LEDD + subtype:sex. subtype: 0 = iPD, 1 = gPD; sex: 0 = female, 1 = male. PD patients only (n = 245). Inference based on heteroscedasticity-consistent robust standard errors (HC3). p (FDR): Benjamini-Hochberg correction applied to the 9 interaction p-values. No interaction term reached significance after FDR correction.

**Table S11**. **Linear model results: subtype × sex interaction on serum NMDAR-related amino acid levels (PD patients only; males as reference category).**

| Outcome | Term | β | SE | t | p (raw) | p (FDR) |
|---|---|---|---|---|---|---|
| **L-Asp** | Intercept | 2.125 | 0.227 | 9.374 | < 0.001 | |
| | **Subtype (gPD vs iPD)** | 0.142 | 0.070 | 2.039 | 0.0425 | 0.0547 |
| | Sex (F vs M) | 0.257 | 0.073 | 3.524 | < 0.001 | |
| | Age | 0.000 | 0.003 | 0.030 | 0.9759 | |
| | Disease duration | -0.002 | 0.004 | -0.491 | 0.6242 | |
| | LEDD | 0.000 | 0.000 | 0.562 | 0.5744 | |
| | **Subtype × Sex (interaction)** | -0.097 | 0.106 | -0.913 | 0.3621 | 0.4073 |
| **L-Glu** | Intercept | 2.851 | 0.312 | 9.146 | < 0.001 | |
| | **Subtype (gPD vs iPD)** | 0.272 | 0.097 | 2.788 | 0.0057 | **0.0129** |
| | Sex (F vs M) | 0.169 | 0.100 | 1.691 | 0.0922 | |
| | Age | 0.001 | 0.004 | 0.244 | 0.8075 | |

| Outcome | Term | β | SE | t | p (raw) | p (FDR) |
|---|---|---|---|---|---|---|
| | Disease duration | -0.004 | 0.006 | -0.622 | 0.5346 | |
| | LEDD | 0.000 | 0.000 | 0.380 | 0.7043 | |
| | **Subtype × Sex (interaction)** | -0.147 | 0.145 | -1.012 | 0.3126 | 0.4019 |
| **L-Asn** | Intercept | 2.463 | 0.322 | 7.643 | < 0.001 | |
| | **Subtype (gPD vs iPD)** | 0.263 | 0.115 | 2.278 | 0.0236 | **0.0425** |
| | Sex (F vs M) | 0.278 | 0.116 | 2.399 | 0.0172 | |
| | Age | 0.002 | 0.005 | 0.334 | 0.7387 | |
| | Disease duration | -0.001 | 0.009 | -0.157 | 0.8755 | |
| | LEDD | 0.000 | 0.000 | 0.382 | 0.7025 | |
| | **Subtype × Sex (interaction)** | -0.209 | 0.165 | -1.267 | 0.2063 | 0.3713 |
| **D-Ser** | Intercept | 0.537 | 0.161 | 3.344 | < 0.001 | |
| | **Subtype (gPD vs iPD)** | 0.207 | 0.063 | 3.312 | 0.0011 | **0.0085** |

| Outcome | Term | β | SE | t | p (raw) | p (FDR) |
|---|---|---|---|---|---|---|
| | Sex (F vs M) | 0.124 | 0.058 | 2.135 | 0.0337 | |
| | Age | 0.004 | 0.002 | 1.926 | 0.0553 | |
| | Disease duration | 0.002 | 0.004 | 0.415 | 0.6784 | |
| | LEDD | 0.000 | 0.000 | 0.664 | 0.5076 | |
| | **Subtype × Sex (interaction)** | -0.135 | 0.090 | -1.493 | 0.1369 | 0.3226 |
| **L-Ser** | Intercept | 3.667 | 0.337 | 10.878 | < 0.001 | |
| | **Subtype (gPD vs iPD)** | 0.332 | 0.117 | 2.839 | 0.0049 | **0.0129** |
| | Sex (F vs M) | 0.386 | 0.115 | 3.366 | < 0.001 | |
| | Age | 0.001 | 0.005 | 0.294 | 0.7691 | |
| | Disease duration | -0.005 | 0.008 | -0.604 | 0.5466 | |
| | LEDD | 0.000 | 0.000 | 0.996 | 0.3204 | |
| | **Subtype × Sex (interaction)** | -0.295 | 0.167 | -1.769 | 0.0782 | 0.3226 |

| Outcome | Term | β | SE | t | p (raw) | p (FDR) |
|---|---|---|---|---|---|---|
| **D-Ser/tot-Ser (ratio)** | Intercept | 1.421 | 0.288 | 4.927 | < 0.001 | |
| | **Subtype (gPD vs iPD)** | -0.114 | 0.097 | -1.185 | 0.2374 | 0.2671 |
| | Sex (F vs M) | -0.245 | 0.097 | -2.542 | 0.0117 | |
| | Age | 0.003 | 0.004 | 0.653 | 0.5144 | |
| | Disease duration | 0.006 | 0.006 | 0.912 | 0.3626 | |
| | LEDD | -0.000 | 0.000 | -0.726 | 0.4684 | |
| | **Subtype × Sex (interaction)** | 0.147 | 0.134 | 1.092 | 0.2760 | 0.4019 |
| **L-Gln** | Intercept | 4.693 | 0.274 | 17.106 | < 0.001 | |
| | **Subtype (gPD vs iPD)** | 0.205 | 0.097 | 2.125 | 0.0346 | 0.0519 |
| | Sex (F vs M) | 0.255 | 0.095 | 2.695 | 0.0075 | |
| | Age | 0.002 | 0.004 | 0.457 | 0.6480 | |
| | Disease duration | -0.000 | 0.007 | -0.019 | 0.9848 | |

| Outcome | Term | β | SE | t | p (raw) | p (FDR) |
|---|---|---|---|---|---|---|
| | LEDD | 0.000 | 0.000 | 0.626 | 0.5317 | |
| | **Subtype × Sex (interaction)** | -0.198 | 0.135 | -1.468 | 0.1434 | 0.3226 |
| **Gly** | Intercept | 4.568 | 0.267 | 17.111 | < 0.001 | |
| | **Subtype (gPD vs iPD)** | 0.270 | 0.086 | 3.144 | 0.0019 | **0.0085** |
| | Sex (F vs M) | 0.403 | 0.090 | 4.501 | < 0.001 | |
| | Age | 0.000 | 0.004 | 0.119 | 0.9051 | |
| | Disease duration | -0.000 | 0.005 | -0.034 | 0.9726 | |
| | LEDD | 0.000 | 0.000 | 1.499 | 0.1353 | |
| | **Subtype × Sex (interaction)** | -0.231 | 0.127 | -1.818 | 0.0703 | 0.3226 |
| **L-Gln/L-Glu (ratio)** | Intercept | 1.841 | 0.204 | 9.010 | < 0.001 | |
| | **Subtype (gPD vs iPD)** | -0.066 | 0.065 | -1.023 | 0.3073 | 0.3073 |
| | Sex (F vs M) | 0.086 | 0.068 | 1.262 | 0.2083 | |

| Outcome | Term | β | SE | t | p (raw) | p (FDR) |
|---|---|---|---|---|---|---|
| | Age | 0.001 | 0.003 | 0.245 | 0.8070 | |
| | Disease duration | 0.004 | 0.004 | 0.869 | 0.3859 | |
| | LEDD | 0.000 | 0.000 | 0.077 | 0.9384 | |
| | **Subtype × Sex (interaction)** | -0.050 | 0.093 | -0.541 | 0.5893 | 0.5893 |

Results from ordinary least squares (OLS) linear models fitted separately for each of the nine amino acid outcomes in PD patients only. Model formula: log(amino acid) ~ subtype + sex + age + disease duration + LEDD + subtype:sex, where subtype = iPD (0) vs gPD (1) and sex = male (0) vs female (1). Coefficients (β), robust standard errors (SE), t-statistics and raw p-values are reported for all model terms. The p (FDR) column reports Benjamini-Hochberg false discovery rate-corrected p-values for two terms: the main effect of subtype, which in this parameterisation reflects the iPD vs gPD difference in male patients (FDR correction across 9 tests); and the subtype × sex interaction term, which tests whether this difference varies significantly between sexes (FDR correction across 9 tests). No interaction term reached significance after FDR correction. All inference is based on heteroscedasticity-consistent (HC3) robust standard errors. n = 245.

**Model:** log(amino acid) ~ subtype + sex + age + disease duration + LEDD + subtype:sex. subtype: 0 = iPD, 1 = gPD; sex: 0 = male (reference), 1 = female. PD patients only (n = 245). Inference based on HC3 robust standard errors. p (FDR) is reported for two terms: **Subtype (gPD vs iPD)** — reflects the iPD vs gPD difference in males; FDR correction across 9 tests. **Subtype × Sex (interaction)** — tests whether the iPD vs gPD difference differs between sexes; FDR correction across 9 tests. Highlighted cells: red = significant subtype effect in males ($p < 0.05$ FDR); blue = significant interaction (none observed).

**Table S12. Demographic and clinical characteristics compared between male and female in both idiopathic PD and genetic PD patients enrolled in the serum collection.**

| PD subtype | Clinical characteristics | Male | | | | Female | | | | $p$ value[a] |
|---|---|---|---|---|---|---|---|---|---|---|
| | | N | Median | IQR | | N | Median | IQR | | |
| Idiopathic PD | Age (years) | 65 | 67,0 | 61,0 | 74,0 | 56 | 69,5 | 62,5 | 74,0 | 0,321 |
| | Age at onset (years) | 65 | 61,0 | 55,0 | 66,0 | 56 | 64,0 | 56,5 | 69,0 | 0,196 |
| | LEDD (mg/die) | 65 | 400,0 | 300,0 | 560,0 | 56 | 400,0 | 302,5 | 600,0 | 0,402 |
| | Disease duration (years) | 65 | 5,0 | 3,0 | 8,0 | 56 | 5,0 | 3,0 | 7,0 | 0,532 |
| | MDS-UPDRS III | 65 | 24,0 | 13,0 | 35,0 | 56 | 21,0 | 13,0 | 28,5 | 0,191 |
| Genetic PD | Age (years) | 64 | 68,0 | 61,5 | 72,0 | 60 | 68,0 | 63,5 | 73,5 | 0,676 |
| | Age at onset (years) | 64 | 60,0 | 55,5 | 65,5 | 60 | 61,5 | 52,5 | 65,0 | 0,556 |
| | LEDD (mg/die) | 64 | 451,0 | 320,0 | 724,0 | 60 | 483,0 | 300,0 | 645,0 | 0,795 |
| | Disease duration (years) | 64 | 6,0 | 3,0 | 9,5 | 60 | 7,0 | 3,5 | 12,0 | 0,151 |
| | MDS-UPDRS III | 64 | 22,5 | 14,5 | 31,0 | 60 | 17,5 | 12,0 | 30,0 | 0,167 |

Abbreviations: N, number of subjects; IQR, interquartile range; LEDD, Levodopa equivalent daily dose; MDS-UPDRS III, Movement Disorders Society Unified Parkinson's Disease Rating Scale, part III.
[a]Mann-Whitney test

**Table S13. Common variants in *GRIN2A* and *GRIN2B* genes encoding NMDAR subunits associated with PD in MNI-cohort and PDGC/UK biobank**

| Sex Cohort (N) | Contrast | Gene | chr | Position hg38 | nt change | position | SNP id | test | p | OR |
|---|---|---|---|---|---|---|---|---|---|---|
| **Male + Female** MNI-PD *vs* MNI-HC (804 *vs* 282) | PD *vs* HC | GRIN2A | 16 | 10112318 | T>C | Intronic | rs11866570 | ADD | NS | NS |
| | | GRIN2B | 12 | 13675725 | T>C | Intronic | rs11055581 | ADD | 0.003 | 1.48 |
| | | GRIN2A | 16 | 10112087 | C>T | Intronic | rs62621078 | ADD | NS | NS |
| | | GRIN2B | 12 | 13865843 | G>C | SV p.P122P | rs7301328 | ADD | NS | NS |
| | | GRIN2B | 12 | 13564574 | G>A | SV p.T888T | rs1806201 | ADD | NS | NS |
| | | GRIN2A | 16 | 9841209 | T>G | Intronic | rs6497540 | ADD | NS | NS |
| **Male + Female** PDGC vs UK-HC (4586 *vs* 43989) | PD *vs* HC | GRIN2A | 16 | 10112318 | T>C | Intronic | rs11866570 | Fisher test | NS | NS |
| | | GRIN2B | 12 | 13675725 | T>C | Intronic | rs11055581 | Fisher test | **< 0.00001** | 1.12 |
| | | GRIN2A | 16 | 10112087 | C>T | Intronic | rs62621078 | Fisher test | NS | NS |
| | | GRIN2B | 12 | 13865843 | G>C | SV p.P122P | rs7301328 | Fisher test | **< 0.00001** | 0.91 |
| | | GRIN2B | 12 | 13564574 | G>A | SV p.T888T | rs1806201 | Fisher test | 0.005964 | 0.94 |
| | | GRIN2A | 16 | 9841209 | T>G | Intronic | rs6497540 | Fisher test | NS | NS |

Abbreviations: N, number of subjects; PD, Parkinson's disease HC, Healthy controls; MNI, Mediterranean Neurological Institute; chr, chromosome; position hg38, genomic position human genome assembly 38; nt change, nucleotide change; SNP id, single nucleotide polymorphism identification number; p, p-value calculated with logistic regression model with Plink software; ADD, additive genetic model; OR, Odds Ratio; SV, synonymous variant; NS, not significant. Significance after Bonferroni correction p=7.1E-04. Genes analysed: *GRIN1, GRIN2A, GRIN2B*

**Table S14. Association analysis of the *GRIN2A* rs11866570 allelic variant in human brain tissues**

| Gene Symbol | Genecode Id | Variant Id | Position Hg38 | P-Value | NES | T-statistic | Tissue |
|---|---|---|---|---|---|---|---|
| GRIN2A | ENSG00000183454 | rs11866570 | 16: 10112318 | 0.69 | 0.022 | 0.40 | Brain - Amygdala |
| GRIN2A | ENSG00000183454 | rs11866570 | 16: 10112318 | 0.81 | -0.0072 | -0.25 | Brain - Anterior cingulate cortex (BA24) |
| GRIN2A | ENSG00000183454 | rs11866570 | 16: 10112318 | **0.00055** | -0.17 | -3.5 | Brain - Caudate (basal ganglia) |
| GRIN2A | ENSG00000183454 | rs11866570 | 16: 10112318 | **0.000028** | 0.26 | 4.3 | Brain - Cerebellum |
| GRIN2A | ENSG00000183454 | rs11866570 | 16: 10112318 | 0.91 | 0.0027 | 0.12 | Brain - Cortex |
| GRIN2A | ENSG00000183454 | rs11866570 | 16: 10112318 | 0.77 | 0.0081 | 0.30 | Brain - Frontal Cortex (BA9) |
| GRIN2A | ENSG00000183454 | rs11866570 | 16: 10112318 | 0.064 | -0.086 | -1.9 | Brain - Hippocampus |
| GRIN2A | ENSG00000183454 | rs11866570 | 16: 10112318 | 0.40 | -0.074 | -0.85 | Brain - Hypothalamus |
| GRIN2A | ENSG00000183454 | rs11866570 | 16: 10112318 | 0.21 | 0.065 | 1.3 | Brain - Nucleus accumbens (basal ganglia) |
| GRIN2A | ENSG00000183454 | rs11866570 | 16: 10112318 | **0.0035** | -0.18 | -3.0 | Brain - Putamen (basal ganglia) |
| GRIN2A | ENSG00000183454 | rs11866570 | 16: 10112318 | 0.98 | 0.0027 | 0.024 | Brain - Substantia nigra |

Genecode Id, id number of the GRIN2A transcript in Ensembl Genome Browser; Variant id, id of the variant in the Single Nucleotide Polymorphism database; Position Hg38, genomic position on Chromosome assembly HG38; NES, enrichment score normalized. Significant p-value are reported in bold

**Table S15. Association analysis of the *GRIN2B* rs7301328 allelic variant in human brain tissues**

| Gene Symbol | Genecode Id | Variant Id | Position Hg38 | P-Value | NES | T-statistic | Tissue |
|---|---|---|---|---|---|---|---|
| GRIN2B | ENSG00000273079 | rs7301328 | 12: 13865843 | 0.96 | 0.0013 | 0.052 | Brain - Amygdala |
| GRIN2B | ENSG00000273079 | rs7301328 | 12: 13865843 | 0.24 | 0.023 | 1.2 | Brain - Anterior qcingulate cortex (BA24) |
| GRIN2B | ENSG00000273079 | rs7301328 | 12: 13865843 | 0.66 | 0.0072 | 0.45 | Brain - Caudate (basal ganglia) |
| GRIN2B | ENSG00000273079 | rs7301328 | 12: 13865843 | 0.60 | -0.014 | -0.52 | Brain - Cerebellum |
| GRIN2B | ENSG00000273079 | rs7301328 | 12: 13865843 | 0.098 | -0.023 | -1.7 | Brain - Cortex |
| GRIN2B | ENSG00000273079 | rs7301328 | 12: 13865843 | 0.29 | -0.021 | -1.1 | Brain - Frontal Cortex (BA9) |
| GRIN2B | ENSG00000273079 | rs7301328 | 12: 13865843 | 0.87 | -0.0027 | -0.16 | Brain - Hippocampus |
| GRIN2B | ENSG00000273079 | rs7301328 | 12: 13865843 | 0.77 | -0.0065 | -0.29 | Brain - Hypothalamus |
| GRIN2B | ENSG00000273079 | rs7301328 | 12: 13865843 | 0.92 | -0.0019 | -0.10 | Brain - Nucleus accumbens (basal ganglia) |
| GRIN2B | ENSG00000273079 | rs7301328 | 12: 13865843 | 0.19 | -0.024 | -1.3 | Brain - Putamen (basal ganglia) |
| GRIN2B | ENSG00000273079 | rs7301328 | 12: 13865843 | **0.0061** | 0.072 | 2.8 | Brain - Substantia nigra |

Genecode Id, id number of the GRIN2A transcript in Ensembl Genome Browser; Variant id, id of the variant in the Single Nucleotide Polymorphism database; Position Hg38, genomic position on Chromosome assembly HG38; NES, enrichment score normalized. Significant p-value are reported in bold

**Table S16.** Association analysis of the *GRIN2B* rs1806201 allelic variant in human brain tissues

| Gene Symbol | Gencode Id | Variant Id | Position Hg38 | P-Value | NES | T-statistic | Tissue |
|---|---|---|---|---|---|---|---|
| GRIN2B | ENSG00000273079 | rs1806201 | 12: 13564574 | **0.00075** | 0.10 | 3.4 | Brain - Amygdala |
| GRIN2B | ENSG00000273079 | rs1806201 | 12: 13564574 | 0.72 | 0.0081 | 0.36 | Brain - Anterior qcingulate cortex (BA24) |
| GRIN2B | ENSG00000273079 | rs1806201 | 12: 13564574 | **0.00039** | 0.065 | 3.6 | Brain - Caudate (basal ganglia) |
| GRIN2B | ENSG00000273079 | rs1806201 | 12: 13564574 | 0.39 | -0.029 | -0.86 | Brain - Cerebellum |
| GRIN2B | ENSG00000273079 | rs1806201 | 12: 13564574 | **0.00017** | 0.061 | 3.8 | Brain - Cortex |
| GRIN2B | ENSG00000273079 | rs1806201 | 12: 13564574 | 0.20 | 0.030 | 1.3 | Brain - Frontal Cortex (BA9) |
| GRIN2B | ENSG00000273079 | rs1806201 | 12: 13564574 | 0.042 | 0.041 | 2.0 | Brain - Hippocampus |
| GRIN2B | ENSG00000273079 | rs1806201 | 12: 13564574 | **0.00064** | 0.092 | 3.5 | Brain - Hypothalamus |
| GRIN2B | ENSG00000273079 | rs1806201 | 12: 13564574 | 0.054 | 0.044 | 1.9 | Brain - Nucleus accumbens (basal ganglia) |
| GRIN2B | ENSG00000273079 | rs1806201 | 12: 13564574 | **0.00013** | 0.084 | 3.9 | Brain - Putamen (basal ganglia) |
| GRIN2B | ENSG00000273079 | rs1806201 | 12: 13564574 | 0.44 | 0.025 | 0.77 | Brain - Substantia nigra |

Genecode Id, id number of the GRIN2A transcript in Ensembl Genome Browser; Variant id, id of the variant in the Single Nucleotide Polymorphism database; Position Hg38, genomic position on Chromosome assembly HG38; NES, enrichment score normalized. Significant p-value are reported in bold

**Checklist**

**1. Reporting Guidelines**
-STROBE Compliance: This study was designed and reported following the Strengthening the Reporting of Observational Studies in Epidemiology (STROBE) guidelines for case-control studies. **Pag. 17 lines 11-14**

-Biochemical Reporting: Biochemical data acquisition and processing comply with the Metabolomics Standards Initiative (MSI) guidelines. **Pag. 17 lines 12-14**

**2. Ethical Compliance & Registration**
**-**Institutional Review Board (IRB): Approved by the IRB of IRCCS Neuromed, Italy (Protocols: N°9/2015, N°19/2020, N°4/2023). **Pag. 17 lines 3-6**

-ClinicalTrials.gov Registration: The study is registered under identifiers NCT02403765, NCT04620980, and NCT05721911. **Pag. 17 lines 3-6**

-Declaration of Helsinki: All clinical investigations were conducted according to the principles of the Declaration of Helsinki. **Pag. 17 lines 7-8**

-Informed Consent: Written informed consent was obtained from all study participants. **Pag. 17 line 8**

**3. Data Stratification & Quality Control**
- Genetic Stratification: Patients were stratified into two distinct groups:
iPD: Idiopathic (no variants in the 37-gene panel). **Pag. 18 lines 21-22**
gPD: Genetic (pathogenic mutations in LRRK2, GBA1, TMEM175, PARK2, PINK1, PARK7). **Pag. 18 lines 23-26; Pag. 19 lines 1-23.**

- Sex-Matching: Healthy controls (HC) were sex-matched to the PD cohort to minimize confounding bias in biochemical analysis. **Pag. 17 lines 22-23**

- Confounder Adjustment: Statistical analyses (ANCOVA and partial correlations) were adjusted for age, disease duration, and L-DOPA Equivalent Daily Dose (LEDD). **Pag. 21 lines 10-14**

**4. Technical Validation**
- HPLC quantification: Amino acid concentrations were quantified via UHPLC with precolumn derivatization. **Pag. 20 lines 9-20**

- Genetic Validation: Key genetic associations (e.g., GRIN2B) were validated in independent larger cohorts (MNI and PDGC/UK). **Pag. 11 lines 9-15**

**5. Data Availability & Reproducibility**
- Software used: PLINK2 (Genetics) **Pag.23 lines 11-12**

- Public Databases: Variant pathogenicity was cross-referenced with LOVD v.3.0 **Pag.19 lines 1-4**. Gene expression data were sourced from the GTEx portal **Pag. 11 lines 19-25**.

**1. Study Design & Setting (STROBE Items 4 & 5)**
- Design Type: Defined as a case-control observational study. **Pag.16 line 3**

- Study Periods: Two recruitment windows specified (June 2015–Dec 2017 and June 2021–Dec 2023) **Pag. 16 lines 7-8.**

- Location: Parkinson Centre of the IRCCS INM Neuromed, Italy **Pag 16. Lines 6-7.**

**2. Participant Selection & Eligibility (STROBE Item 6)**
-Case Definition: PD diagnosis based on ≥2 cardinal motor signs (tremor, bradykinesia, rigidity) and positive response to L-DOPA. **Pag. 16 lines 12-14**

-Control Definition: Healthy subjects (HC) negative for PD gene mutations, matched for sex with the PD cohort. **Pag. 19 lines 24-25 and Pag.17 line 23.**

-Age Threshold: Inclusion limited to individuals aged ≥40 years to maintain cohort relevance. **Pag.16 lines 5-6**

- Exclusion Criteria: Explicitly listed (pre-existing psychiatric conditions, other neurodegenerative diseases like MS or ALS, dementia, depression, and use of specific psychotropic medications). **Pag. 16 lines 15-19**

**3. Data Sources & Clinical Assessment (STROBE Item 8)**
-Clinical Scale: MDS-UPDRS Part III used for motor symptom severity (assessed during the "ON" period). **Pag. 16 line 23.**

-Ancestry: Confirmed European ancestry for all participants. **Pag. 16 lines 6-7**

-Biobank Origin: Subjects selected from the IRCCS Neuromed/IGB-CNR biobank. **Pag.16 lines 4-5**

**4. Genetic Stratification Framework**
- Panel Composition: Use of a 37-gene panel (10 Mendelian PD genes + 27 risk factor genes). **Pag. 18 lines 11-16.**
- Sequencing Method: Whole Exome Sequencing (WES) data analyzed for rare exonic variants (MAF < 0.01 based on gnomAD v.4.1.0). **Pag.18 lines 9-21.**

- Group Classification:
iPD (Idiopathic): No mutations in the 37-gene panel. **Pag. 18 lines 21-23.**
gPD (Genetic): Carrying known pathogenic mutations (e.g., LRRK2 G2019S, GBA1). **Pag. 18 lines 24-26; Pag. 19 lines 1-24.**

**5. Laboratory & Analytical Protocols**

- Serum Handling: Standardized 6-hour fasting collection, 30-min clotting, and -80°C storage. **Pag. 20 line 2-8.**

- HPLC Parameters: Methanol dilution **Pag.20 lines 11-3**, TCA neutralization **Pag.20 lines 13-14**, precolumn derivatization (OPA/NAC) **Pag.20 lines 13-14**, and C18 reversed-phase column separation **Pag.20 lines 14-17.**

- Standardization: Use of anonymized codes and internal reference signals (external standards) **Pag. 20 lines 7-8 and Pag. 20 lines 19-20**.

**6. Statistical & Confounding Control (STROBE Items 10 & 12)**
- Sample Size: Acknowledged as determined by biobank availability (no formal a priori calculation). **Pag. 17 lines 13-14**

- Confounder Adjustment: Use of ANCOVA for age, LEDD, and disease duration; Natural log transformation for non-normal distributions. **Pag. 21 lines 6-8**

- Multiple Testing: Bonferroni correction applied to p-values in genetic association tests. **Pag. 11 lines 7-8; Pag. 23 lines 13-14**
